# The Subcortical Default Mode Network and Alzheimer’s Disease: A systematic review and Activation Likelihood Estimation Meta-Analysis

**DOI:** 10.1101/2023.10.02.23296420

**Authors:** Sara L Seoane, Martijn van den Heuvel, Ángel Acebes, Niels Janssen

## Abstract

The default mode network is a central cortical brain network suggested to play a major role in several disorders, and to be particularly vulnerable to the neuropathological hallmarks of Alzheimer’s disease. Subcortical involvement in the default mode network and its alteration in Alzhimer’s disease remains largely unknown. We performed a systematic review, meta-analysis, and empirical validation of the subcortical default mode network in healthy adults, combined with a systematic review, meta-analysis, and network analysis of the involvement of subcortical default mode areas in Alzheimer’s disease. Our results show that, besides the well-known cortical default mode network brain regions, the default mode network consistently includes subcortical regions, namely the thalamus, lobule and vermis IX and right Crus I/II of the cerebellum, and the amygdala. Network analysis also suggests the involvement of the caudate nucleus. In Alzheimer’s disease, we observed a left-lateralized cluster of decrease in functional connectivity which covered the medial temporal lobe and amygdala and showed overlap with the default mode network in a portion covering parts of the left anterior hippocampus and left amygdala. An increase in functional connectivity was also found in the right anterior insula. These results confirm the consistency of subcortical contributions to the default mode network in healthy adults and highlight the relevance of the subcortical default mode network alteration in Alzheimer’s disease.

## 1 Introduction

Our understanding of the default mode network emerged from the observation of a characteristic spatial pattern within the brain of increased blood flow during resting wakefulness and decreased blood flow during externally oriented tasks [1, 2]. The default mode network is indeed considered the most prevalent brain spatial pattern of coactivation during resting state [3–7], and a network crucially implicated in mental and neurological disorders such as schizophrenia [8], multiple sclerosis [9], and Alzheimer’s disease [10, 11]. Our understanding of its cortical components has greatly advanced [12–16]. However, the study of the subcortical components and their alterations, particularly in the context of Alzheimer’s disease, has not yet derived a clear list of subcortical structures. This study aims to address this lacuna, systematically investigating the subcortical regions within the default mode network and their functional connectivity alterations in Alzheimer’s disease.

Previous studies identified subcortical brain regions within the default mode network [17–20]. The amygdala is reported as part of the pattern of decreased blood flow during tasks in a hallmark meta-analysis of positron emission tomography (PET) that was crucial in forming what we recognize today as the default mode network [2]. An earlier work on the brain activity mapping of the awake rest, also using PET, found the cerebellum as part of this characteristic resting-state pattern [21]. An increasing number of recent studies find subcortical functional and anatomical connections to the default mode network in subcortical regions that include the amygdala, anterior and mediodorsal thalamus, basal forebrain, nucleus accumbens, medial septal nucleus, ventral tegmental area, caudate nucleus, and hypothalamus [3, 19, 22–28].

Given the known impact of Alzheimer’s disease on subcortical structures [29–33], as well as the relevance of the default mode network to pathological protein aggregation in the disease [11, 33, 34], our study aims to identify the subcortical regions of default mode network alterations in Alzheimer’s disease. Previous studies have mainly observed decreased functional connectivity in the default mode network in Alzheimer’s disease [35], and increases were found when looking at subnetworks within the default mode network [36, 37]. The hippocampus, a key component of the default mode network which is especially vulnerable to damage at the early stages of Alzheimer’s disease [38], displays both increases and decreases in its functional connectivity with the rest of the brain and with the default mode network in particular [39–41]. The amygdala, the thalamus, and Crus II and lobule IX of the cerebellum have shown decreased functional connectivity to default mode network regions [31, 32, 42].

Regardless of these advances made, our understanding of the subcortical components of the default mode network as well as of the subcortical default mode network alterations in Alzheimer’s disease remains incomplete without consistent results, which may be linked to at least three main reasons. First, a branch of neuroimaging analysis methods based on the projection of the volume cortical data to a surface has improved our cortical spatial precision [43] but often overlooks subcortical regions and the hippocampus if not used in combination with volume-based methods [44]. Second, the signal-to-noise ratio is lower in some areas of the brain, including subcortical structures [45, 46]. Given that the detection of functional connectivity between brain regions is influenced by the quality of the signals [22], the regional difference in signal quality could impact our ability to detect functional connectivity. Third, subcortical structures are small and variable, which makes it difficult to align them for group analysis [18]. Thus, a systematic study of the findings of subcortical default mode network components and their alterations in Alzheimer’s disease is a crucial task not yet done.

In this study, we performed a systematic review, meta-analysis, and empirical validation of the cortical and subcortical default mode network brain regions in healthy individuals. We also conducted a systematic review, meta-analysis, seed-based network analysis, and conjunction analysis of the cortical and subcortical brain sites that present functional connectivity alterations in Alzheimer’s disease and their correspondence with default mode network brain regions (see Table 1). Provided the aforementioned methodological limitations we reduced bias in examining alterations of the subcortical default mode network in Alzheimer’s disease by meta-analyzing the cortical and subcortical brain sites that show altered functional connectivity in general in the disease, and then determining whether these brain sites were part of the default mode network. Given previous research, we expected to find subcortical structures of the default mode network including the thalamus, the caudate nucleus, the amygdala, the Crus I/II and lobule IX of the cerebellum, and the basal forebrain. Another anticipation had been to identify brain cortical and subcortical sites of decreased functional connectivity in Alzheimer’s disease that included the precuneus, hippocampus, and thalamus. Identifying the consistent involvement of subcortical structures in the default mode network and the changes in functional connectivity in Alzheimer’s disease has the potential to impact our knowledge of the network’s role in the disease.

**Table 1.** Summary of PICO (Population, Intervention, Comparator, and Outcome) components per question.

|  | **Default mode network in healthy participants** | **FC alterations and their directions in AD** |
| --- | --- | --- |
| **Population** | Healthy participants. | Patients that meet the criteria for possible or probable AD. |
| **Intervention** | FC analysis of the default mode network or its seeds performed on whole-brain data. | Comparison of whole-brain FC between AD and matched HC groups (AD > HC and AD < HC). |
| **Comparator** | None needed. | Healthy elderly controls. |
| **Outcome** | What are the cortical and subcortical nodes of the default mode network? | What are the brain sites of increases and decreases in FC in AD? How do they relate to the default mode network? |

**Abbreviations:** AD, Alzheimer’s dementia; HC, healthy controls; FC, functional connectivity.

## 2 Materials and Methods

### 2.1 Search strategy and database selection

This study follows the Preferred Reporting Items for Systematic Reviews and Meta-Analyses (PRISMA) guidelines [47] and uses PubMed, Scopus, Web of Science (WoS), and NeuroVault [48] for database searches. The search was conducted between May 4, 2022, and June 1, 2022, and included the “All Fields” option in PubMed, title, abstract, and keywords in Scopus, and title, abstract, keywords, and automatically generated terms from the titles of the cited papers in Web of Science. We did not limit our search to a specific timeframe. The search details, including queries, search dates, and number of results are documented in Supplementary Tables 1 and 2. We also scrutinized the reference sections and the studies previously known to the authors to ensure comprehensive eligibility consideration (see Fig. 1).

**Table 2.** Clusters coverage in the resting-state default mode network in healthy adults.

| Cluster number | Brain regions covered | Lateralization | Coordinates in MNI152 space (peak ALE value) | ALE | P-value | Z-value |
| --- | --- | --- | --- | --- | --- | --- |
| 1 | Posterior cingulate gyrus, precuneus | Bilateral (65.2% L; 34.8% R) | -4 -56 22 | 0.083 | 2.29E-19 | 8.92 |
| 2 | Inferior parietal lobule, angular gyrus | Left (100%L) | -48 -66 34 | 0.081 | 9.65E-19 | 8.76 |
| 3 | Medial frontal pole | Bilateral (55.5% L; 44.5% R) | 8 42 -6 | 0.050 | 3.88E-10 | 6.15 |
| 4 | Parahippocampal gyrus, hippocampus, amygdala | Right (100% R) | 28 -14 -20 | 0.050 | 2.56E-10 | 6.22 |
| 5 | Inferior parietal lobule, angular gyrus | Right (100% R) | 52 -60 34 | 0.062 | 2.45E-13 | 7.23 |
| 6 | Parahippocampal gyrus, hippocampus, amygdala | Left (100% L) | -28 -24 -12 | 0.048 | 1.25E-9 | 5.96 |
| 7 | Thalamus | Bilateral (62.8% L; 37.2% R) | -2 -12 6 | 0.051 | 2.14E-10 | 6.24 |
| 8 | Crus I/II of the cerebellum (pyramis, inferior semilunar lobule, and uvula) | Right (100% R) | 30 -78 -34 | 0.037 | 3.52E-7 | 4.96 |
| 9 | Lobule and vermis IX of the cerebellum | Bilateral (76.7% L; 23.3% R) | -6 -56 -46 | 0.040 | 7.92E-8 | 5.24 |
| 10 | Ventromedial prefrontal cortex, anterior cingulate cortex | Bilateral (87.9% L; 12.1% R) | 0 50 32 | 0.042 | 2.41E-8 | 5.46 |

**Abbreviations:** ALE, activation likelihood estimation; dlPFC, dorsal lateral prefrontal cortex; L, left; MNI, Montreal Neurological Institute; R, right; SMA, supplementary motor area.

**Fig. 1.**
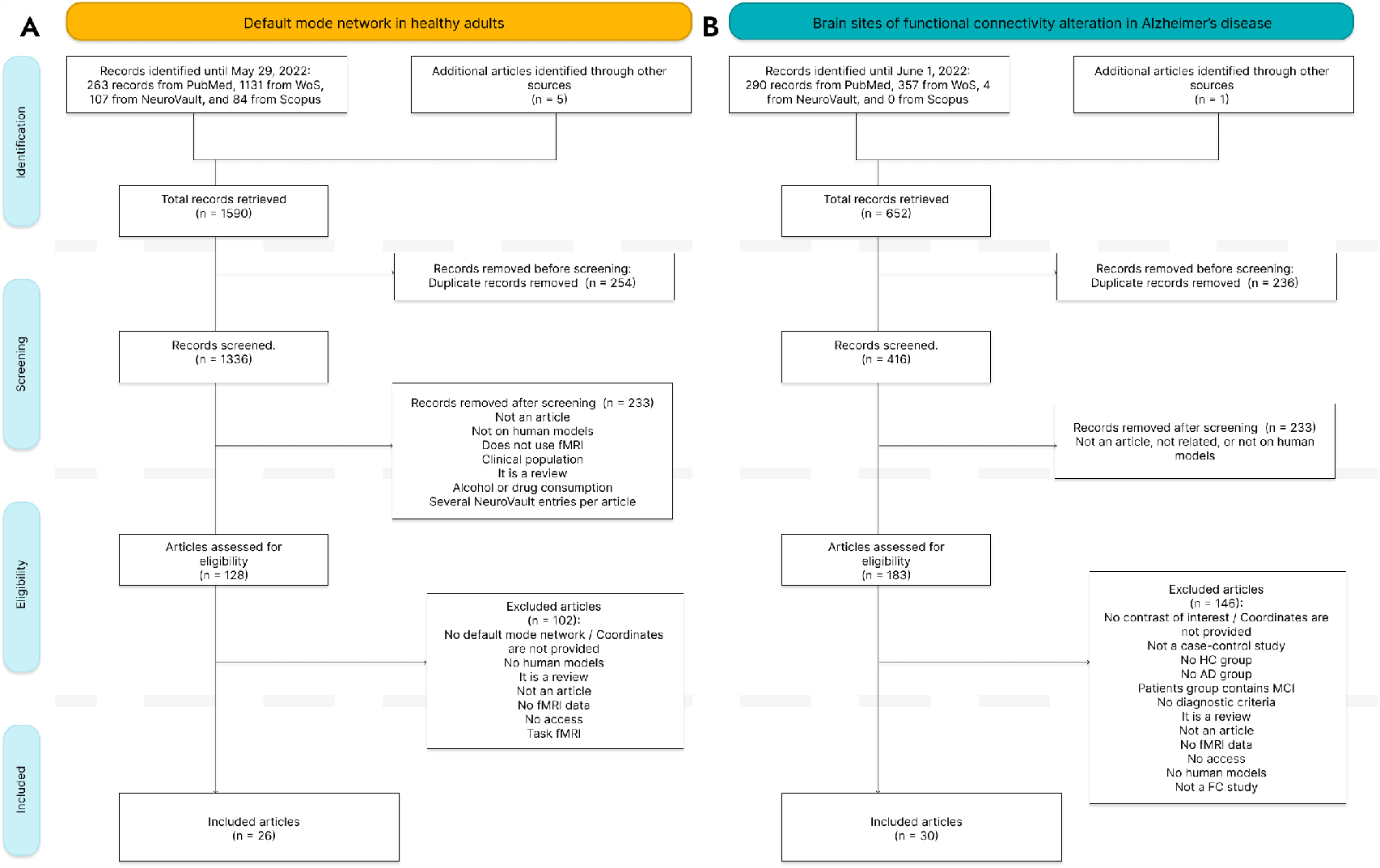
PRISMA workflows. (A) Workflows for questions 1 (B) and 2. The workflow is divided into four phases (from top to bottom): identification, screening, eligibility, and inclusion. In the identification phase, the initial search retrieved 1,586 records for question 1, and 652 records for question 2. These records were reduced in the screening and eligibility phases to 26 and 30 articles that were used in this meta-analysis.

### 2.2 Selection criteria

Study search and selection of studies were performed by SLS and revised by NJ. For the meta-analysis of the default mode network in healthy adults, included studies were whole-brain studies that examined the intrinsic functional connectivity of the default mode network in healthy adults and which reported results in standard anatomical space (MNI or Talairach). The selected studies used samples of 15 or more adult human participants aged 18 - 55 years old. Only studies that examined fMRI data from the whole brain (using volume-based or combined surface- and volume-based methods) were considered. This excluded, for example, studies with reduced fields of view or that only included cortical regions in their analysis. Coordinates that were reported in the manuscripts as part of the default mode network, or connected to its most remarkable brain regions (i.e., the precuneus/posterior cingulate cortex/retrosplenial cortex) were included in the analysis. Negative functional connectivity or “anticorrelations” were not considered for the analysis. Resting-state fMRI studies were considered for inclusion, regardless of whether they had closed or open eyes and whether there was a visual fixation.

For the meta-analysis of brain sites of coactivation alteration in Alzheimer’s disease, included studies were case-control studies that examined whole-brain functional connectivity differences between Alzheimer’s disease (AD) patients and healthy controls (HC). Only records in which the Alzheimer’s disease patients met the criteria for possible or probable Alzheimer’s disease published by the National Institute of Neurological and Communicative Diseases and Stroke/Alzheimer’s Disease and Related Disorders Association (NINCDS-ADRDA) or the National Institute on Aging-Alzheimer’s Association (NIA-AA) were included. All included studies had at least 10 participants in both patients and control groups. All experiments from the selected articles that consisted of AD *>* HC or AD *<* HC contrasts were included. Finally, the quality of the studies included in the meta-analysis of Alzheimer’s disease brain sites was assessed using the Critical Appraisal Skills Programme (CASP [49]) checklist for case-control studies.

### 2.3 Data extraction

For the meta-analysis of the default mode network in healthy adults, the extracted data included sample size, age, gender proportion, network extraction technique, coordinate space in which the coordinates are provided in the study, presence of subcortical regions in the results, and coordinates of the default mode network (see Supplementary Table 3). Variables extracted from the included studies in the meta-analysis of Alzheimer’s disease altered brain sites were sample size, gender proportion, and age of Alzheimer’s disease and healthy control groups, diagnostic criteria for the Alzheimer’s disease, main analysis technique used, and coordinates for brain sites with altered functional connectivity in Alzheimer’s disease compared to healthy controls (see Supplementary Tables 4 and 5).

The extracted data included sets of coordinates, or foci, in either MNI or Talairach space. These foci were extracted from the manuscripts, supplementary materials, and openly available brain images. Each included article could have multiple functional connectivity maps for the default mode network or sets of brain sites of functional connectivity alteration in Alzheimer’s disease. Each functional connectivity or contrast map was treated as an independent experiment in the meta-analysis. See more in the Supplementary methods.

### 2.4 Meta-analysis procedure

Three meta-analyses of brain maps were performed. First, a meta-analysis using foci from experiments of the default mode network in healthy adults. Second, a meta-analysis using foci with increased functional connectivity in Alzheimer’s disease patients against healthy controls (AD *>* HC). And third, a meta-analysis of foci with decreased functional connectivity in Alzheimer’s disease patients against health controls (AD *<* HC). Separate foci files were prepared for the three meta-analyses, and Activation Likelihood Estimation (ALE) was calculated using the GingerALE software [50, 51]. ALE meta-analysis is a coordinate-, and kernel-based method for meta-analysis of neuroimaging data. This method consists of three main steps. First, the interpolation of a three-dimensional Gaussian distribution (or kernel) around each focus (set of 3 coordinates). Second, a combination of the resulting maps for each experiment. Third, the calculation, through a non-additive random effects model, of a single combined map that represents the consistent activations across experiments. This model uses the sample size per experiment, given its impact on each experiment’s power, to find statistical effects. Moreover, the significance of this map is assessed by a Monte Carlo test under the null hypothesis of complete spatial randomness [52–54]. The three meta-analyses were performed using the default cluster-level family-wise estimation (FWE) correction at *p <* 0.01, 1000 permutations, and thresholding of *p <* 0.001.

### 2.5 Data set used for empirical validation and network analysis

A resting-state fMRI sample was used in both the validation analysis of the default mode network brain regions and the network analysis from the altered brain sites in Alzheimer’s disease. This dataset consisted of resting-state fMRI data from 172 healthy young participants in the 7T WU-Minn Human Connectome Project’s 1200 subjects data set. These participants were between 22 and 35 years old, and 60% were female. Please find more information on the study participants elsewhere [55]. See more in the supplementary methods. The data analyses were conducted in agreement with the declaration of Helsinki and with the protocol established by the Ethics Commission for Research of the Universidad de La Laguna, the Comité de Ética de la Investigación y Bienestar Animal.

### 2.6 Empirical validation of the cortical and subcortical default mode network

The brain regions identified in the meta-analysis of the default mode network in healthy adults served as seeds to calculate new functional connectivity maps. Spherical regions of interest (ROIs) were created around the peak ALE MNI coordinates of the brain regions resulting from the meta-analysis of the default mode network in healthy adults using the fslmaths program in FSL. The spheres had a radius of 4 mm each, and the average time series of the voxels inside each spherical ROI was extracted for each subject and resting state session using the fslmeants program in FSL. Each average time series was regressed to the individual whole-brain rs-fMRI data in a general linear model with fsl glm program. Group-level functional connectivity analysis was performed for each ROI using FSL randomise program and consisted of randomized non-parametric voxel-wise one-sample t-tests (5000 permutations), threshold-free cluster enhancement, and *p <* 0.001 as the statistical threshold. A conjunction analysis of the group functional connectivity maps was performed using the AFNI function 3dcalc to determine the overlap between the functional connectivity maps of individual ROIs. An additional seed-based functional connectivity analysis was performed using the same procedure as with each individual ROI, but regressing the average time series across the ten seeds. A group-level functional connectivity map was also achieved using randomise with randomized non-parametric voxel-wise one-sample t-tests (5000 permutations), threshold-free cluster enhancement, and *p <* 0.001 as the statistical threshold.

### 2.7 Network analysis of functional connectivity alterations in Alzheimer’s disease

The connection between the meta-analytical brain sites of functional connectivity alteration in Alzheimer’s disease and the default mode was assessed through a seed-based analysis following the same procedure as per the empirical validation of the default mode network brain regions, except that this time no conjunction analysis was performed.

## 3. Results

### 3.1 Filtered search results

Systematic searches were performed separately for published studies related to the default mode network resting-state functional connectivity in healthy adults, and to the functional connectivity differences between Alzheimer’s disease patients and healthy controls. The two independent searches retrieved 1,585 and 651 records respectively. Six additional articles were identified and included in the lists, for a total of 1,590, and 652 records. After the removal of duplicate records, a total of 1,336 records remained of the searches of the default mode network in healthy adults, and a total of 417 of the searches of the Alzheimer’s disease functional connectivity differences. The first screening was performed to filter records that were not related to the questions, records that were not human model studies, that did not use fMRI, that studied clinical populations, or that were not scientific research articles. 128 articles of the default mode network in healthy adults, and 183 articles of the Alzheimer’s disease functional connectivity differences were assessed for eligibility, and 26 and 30 studies were included in the meta-analyses (see Fig. 1).

A total of 852 foci from 55 experiments in 26 articles were used to calculate a meta-analytical restingstate default mode network. The sample sizes ranged from 15 to 500 (median N = 59), with a weighted pooled proportion (pP) of 49% male participants, and a weighted pooled mean (pM) age of 29.28 years old (range 20.63 - 42.30 years old). The total sample size across all these experiments was 5,165.

For the AD *<* HC contrast of the Alzheimer’s disease brain sites studies, 40 experiments from 26 studies were meta-analyzed. For each experiment, the smallest sample size between case and control groups was chosen for the neuroimaging meta-analysis, leading to a total sample size of 927. The number of Alzheimer’s disease patients per experiment ranged from 10 to 70 (median = 20.71), pP = 45% male, and the number of healthy controls ranged from 10 to 174 (median = 32.88), with a pP = 44% male. Alzheimer’s disease participants had a pM of 71.03 years old, and healthy control participants were 68.34 years old on pM. For the AD *>* HC contrast, 20 experiments from 15 studies were meta-analyzed. Sample sizes in the Alzheimer’s disease group ranged from 10 to 70 (median = 19), pM age = 73.69 years old, and pP male = 46%. Sample sizes in the healthy control groups ranged from 10 to 67 (median = 16.50), pM age = 70.68, and pP male = 48%. Again taking into account the smallest sample size between cases and controls, there was a total sample size of 426 for the meta-analysis of neuroimaging data.

### 3.2 Cortical and subcortical components of the default mode network

The ALE meta-analysis of resting-state experiments of the default mode network in healthy participants resulted in 10 consistent clusters. These clusters covered the (extensively documented) cortical posterior and anterior cingulate cortices, precuneus, inferior parietal lobule, angular gyrus, medial frontal pole, ventromedial prefrontal cortex, anterior and middle parahippocampal gyrus, and hippocampus. Regarding the main focus of this study, the clusters also covered the subcortical thalamus, amygdala, and Crus I/II and lobule and vermis IX of the cerebellum (see Table 2 and Fig. 2 [56]). Contributions from cortical regions, thalamus, and cerebellar lobule IX to the default mode network were bilateral, and cerebellar Crus I/II contributions were right-lateralized.

**Fig. 2.**
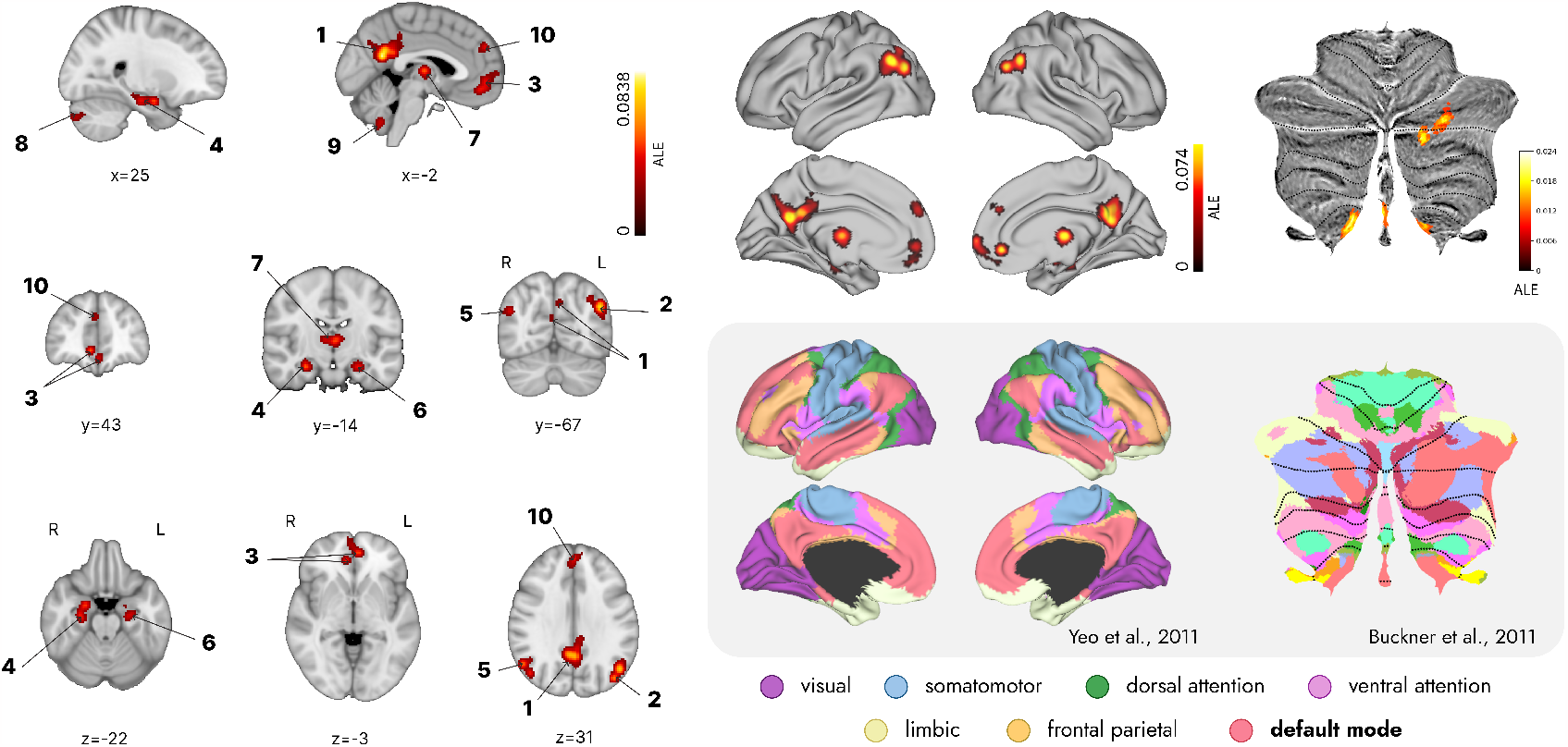
Cortical and subcortical brain regions of the default mode network. (A) The left panel displays the 10 spotted cortical and subcortical clusters of the resting-state default mode network on the standard MNI152 volume. (B) In the top right, the same clusters are projected to a cortical surface (D) and to a cerebellar flatmap for comparison with the default mode network represented in red colors on panels C and E. (C) In the bottom right, the Yeo et al. (2011) 7 resting-state network parcellation is displayed on a cortical surface (E) and Buckner et al. (2012) cerebellar network parcellation is displayed on a cerebellar flatmap.

### 3.3 Brain sites of functional connectivity alterations in Alzheimer’s disease

One cluster of functional connectivity decrease and another cluster of functional connectivity increase was found in Alzheimer’s disease compared to controls (see Fig. 3). The brain site of decreased coactivation was 100% left-lateralized and covered the parahippocampal gyrus, amygdala, and hippocampus. It had a maximum ALE value at -26, -8, and -26 MNI coordinates (*ALE* = 0.023; *p − value* = 8.412384*E −* 7; *z − value* = 4.79). The cluster of increased coactivation was 100% right-lateralized and covered the anterior insula and very little of the precentral gyrus, with a maximum ALE value of 0.0164 at 42, 14, and -2 MNI coordinates (*p − value* = 3.911245*E −* 6; *z − value* = 4.47).

**Fig. 3.**
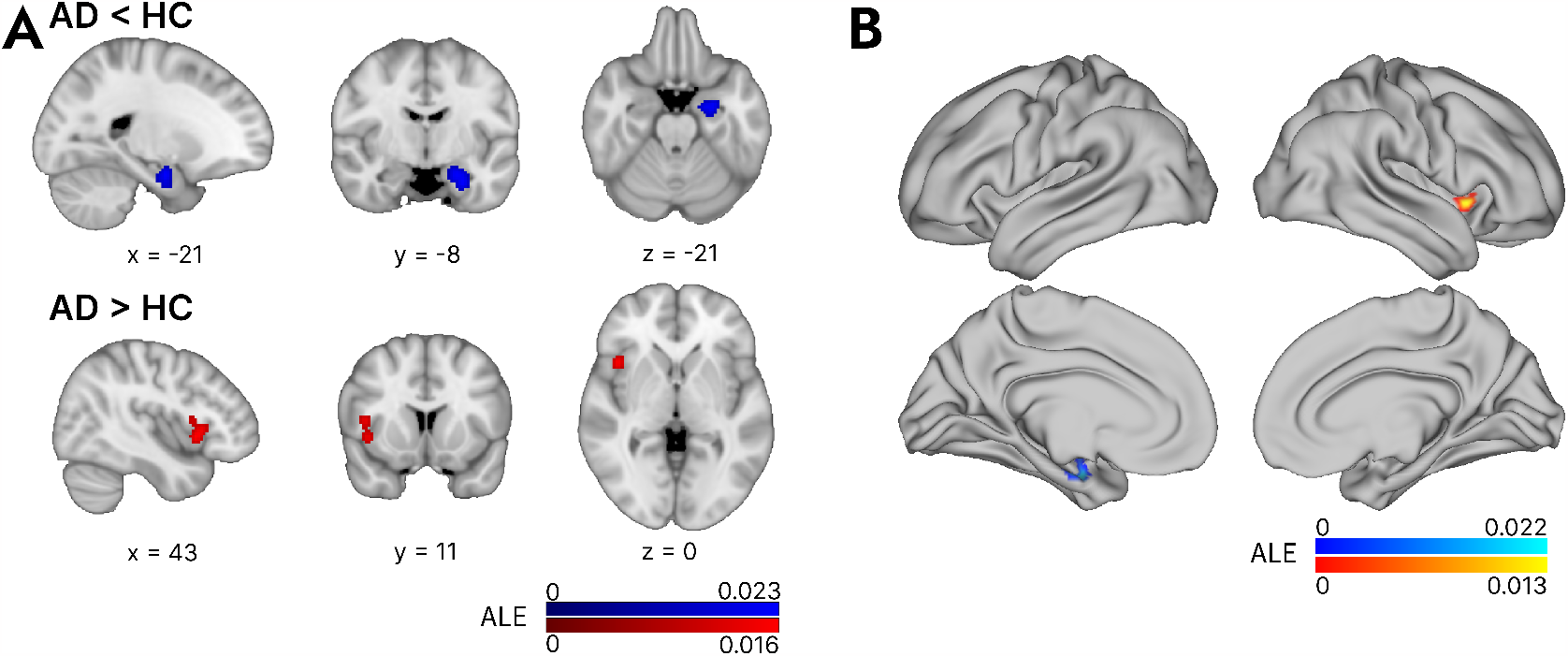
Brain sites of functional connectivity alterations in Alzheimer’s disease. (A) This figure shows a volume view in standard MNI space of the Activation Likelihood Estimation clusters with increased and decreased functional connectivity in Alzheimer’s disease. The cluster in red indicates increases in Alzheimer’s disease against controls, and the cluster in blue indicates decreases. (B) In b, the same clusters are projected to the average surface for comparison with the resting-state networks displayed in Figure 1A.

### 3.4 Empirical validation of the default mode network clusters

The group map of functional connectivity to all ROIs displayed coactivation with precuneus, posterior cingulate cortex, ventromedial prefrontal cortex, frontal pole, orbitofrontal cortex, anterior insula, inferior parietal cortex, precentral and postcentral gyri, hippocampus, left middle temporal gyrus, left fusiform gyrus, bilateral Crus I/II and in lobule IX of the cerebellum, mediodorsal and pulvinar nuclei of the thalamus, caudate nucleus, amygdala, basal forebrain (see Fig. 4A and Supplementary Table 6). Each meta-analytical seed showed significant functional connectivity to the other default mode network brain regions (see Supplementary Figures 1 and 2 and Supplementary Tables 7 to 17). The conjunction analysis of functional connectivity maps of meta-analytical ROIs showed that all seeds connected to the precuneus, posterior cingulate cortex, ventromedial prefrontal cortex, frontal pole, inferior parietal cortex, hippocampus, left middle temporal gyrus, and small clusters in the parahippocampal cortex, bilateral Crus I/II and in lobule IX of the cerebellum, anterior insula, basal forebrain, and in the pulvinar nuclei of the thalamus (see Fig. 4B).

**Fig. 4.**
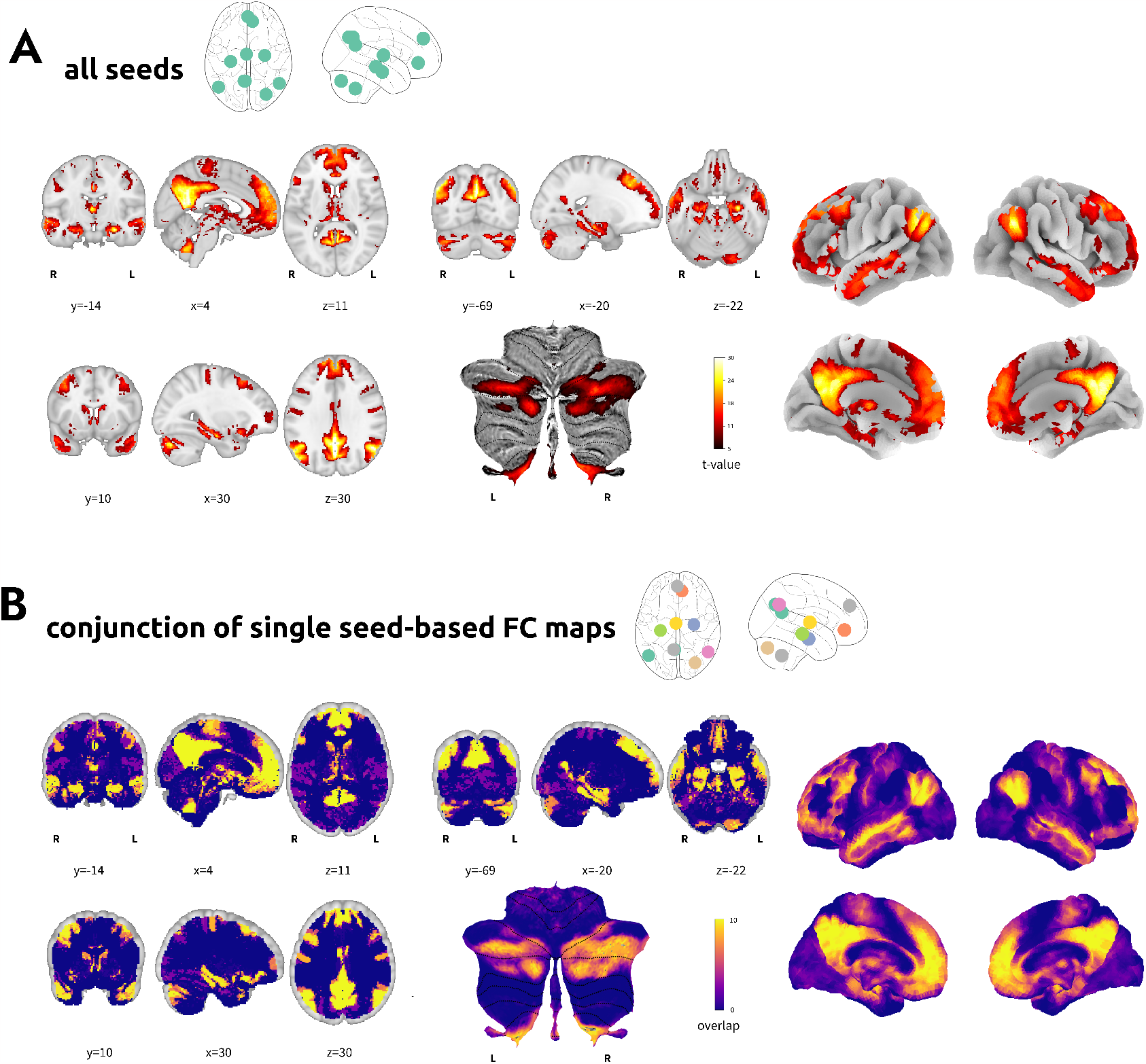
Validation analysis of the default mode network in the HCP 7T dataset. (A) A group functional connectivity map is displayed in panel a for the functional connectivity map to the mask containing all clusters. (B) Panel b shows results from the conjunction analysis of the functional connectivity maps from each seeded cluster (see Supplementary Figures 1 and 2). All maps are displayed in volume, on a surface, and on a cerebellar flat surface for visual comparison with other studies.

### 3.5 Networks of the brain sites of functional connectivity alterations in Alzheimer’s disease

The seed-based functional connectivity analysis of the brain site of decreased functional connectivity in Alzheimer’s disease showed connectivity to the precuneus cortex, posterior cingulate cortex, ventromedial prefrontal cortex, precentral and postcentral gyri, bilateral hippocampi, amygdala, parahippocampal gyri, superior and middle temporal gyri, temporal pole, inferior parietal cortex, basal forebrain, Crus I/II and lobule IX of the cerebellum, and fusiform cortex (see Fig. 5A). The brain site of functional connectivity increase displayed connectivity to the cuneus, intracalcarine and supracalcarine cortex, lingual gyrus, occipital pole, insula, opercular cortex, inferior parietal lobule, intraparietal sulcus, anterior and posterior cingulate cortices, supplementary motor cortex, precentral and poscentral gyri, ventromedial, orbitofrontal and dorsolateral prefrontal cortices, triangularis and opercularis, lobules V, VI, VIIb, VIIIa, IX and Crus II of the cerebellum, putamen, and middle thalamus (see Fig. 5B).

**Fig. 5.**
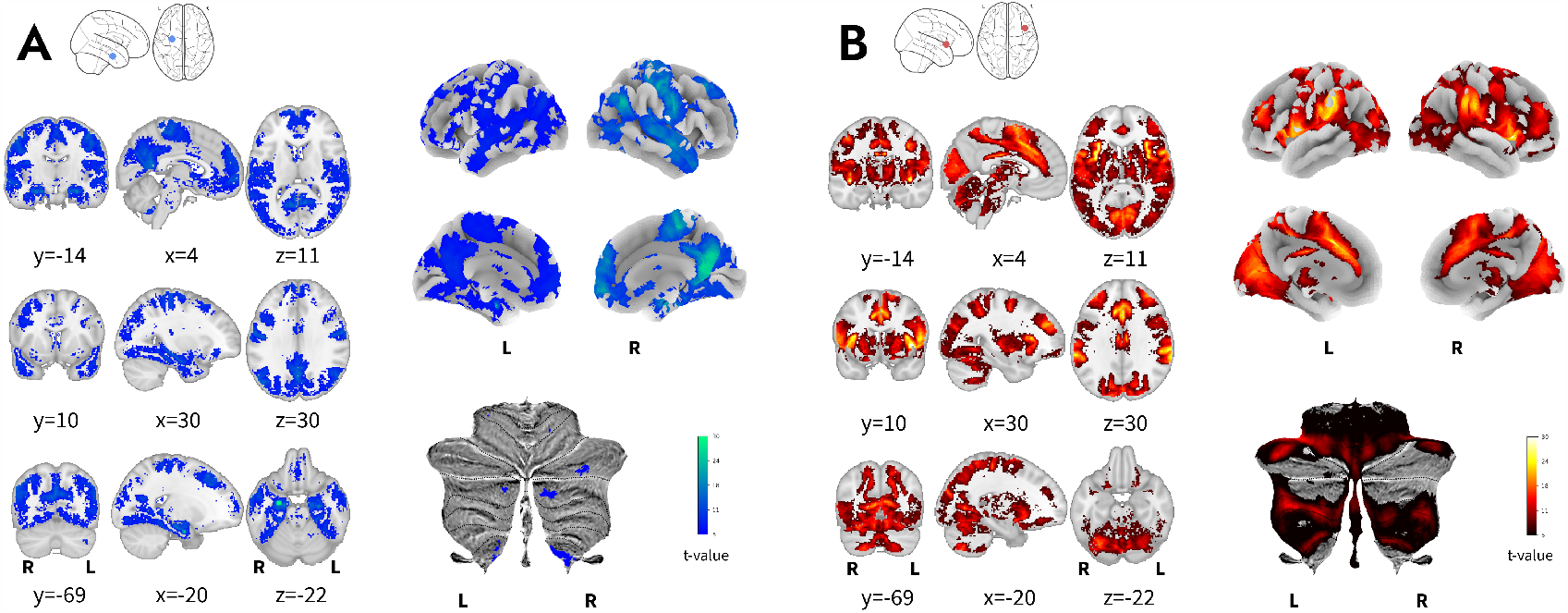
ROI-to-network analysis of Alzheimer’s disease altered brain sites. (A) Panel a shows the functional connectivity map from the ALE meta-analysis cluster of decreased functional connectivity. (B) Panel b shows the functional connectivity map from the meta-analysis cluster of increased connectivity.

### 3.6 Conjunction analysis of default mode network and Alzheimer’s disease alterations

Given the meta-analytical clusters for the default mode network and the functional connectivity alteration in Alzheimer’s disease, we observed an overlap between the default mode network’s cluster 6 and the AD cluster of decreased functional connectivity. To extract the common cluster of voxels between these two clusters we thresholded the volumes with ALE values of the default mode network meta-analysis and the meta-analysis of decreased functional connectivity in Alzheimer’s disease at *ALE* = 0.015 and performed a conjunction analysis, again using the AFNI function 3dcalc. The resulting overlapping cluster consisted of 902,629 voxels that covered the left anterior hippocampus and left amygdala around MNI coordinates -22, -9, and -21.

## 4 Discussion

The default mode network has been extensively studied in the cortex and increasingly studied in the subcortex. Being results in the previous literature variable, a definitive understanding of which subcortical regions are part of the default mode network and exhibit functional connectivity alteration in Alzheimer’s disease has yet to be achieved. Our study combined systematic review, meta-analysis, empirical validation, and network analysis to examine the whole-brain intrinsic functional connectivity maps reported as part of the default mode network in healthy adults and those showing altered functional connectivity in Alzheimer’s disease. We identified and validated ten clusters within the default mode network of healthy adults, seven of which were cortical (two partially subcortical) and three subcortical. These findings, together with our finding of a consistent decrease in functional connectivity in the left anterior hippocampus and left amygdala overlapping with the subcortical default mode network, expand our understanding of the default mode network and its relevance to Alzheimer’s disease.

From the ten clusters identified in the default mode network, seven clusters were located in typical default mode network cortical regions, namely the precuneus and posterior cingulate cortex, the inferior parietal lobule, angular gyrus, ventromedial prefrontal and anterior cingulate cortices, medial frontal pole and the medial temporal lobe, including the hippocampus [57]. The validation analysis revealed functional connectivity between the meta-analytical default mode network brain regions and additional cortical brain regions of the default mode network: the middle and superior temporal gyri and the dorsal prefrontal cortex [57].

Our results extend beyond well-known cortical brain regions, revealing the consistent inclusion of subcortical brain regions in the default mode network. The subcortical brain regions included the thalamus, the amygdala, and specific areas of the cerebellum - crus I/II and lobule and vermis IX, which is consistent with previous results of volume-based and combined surface- and volume-based studies of the default mode network. This study supports the functional connectivity of the thalamus with the default mode network, a finding in various prior studies [18, 19, 58–64]. Our meta-analysis highlights a middle and medial cluster covering the mediodorsal nucleus, and our validation analysis points to a medial and posterior thalamic cluster covering the pulvinar complex. Previous studies find functional connectivity between the default mode network and the anterior mediodorsal thalamus [18], the central lateral nucleus [19], the paraventricular thalamus [60, 62], and to an anterior portion of the right and a posterior portion of the left thalamus [65]. Given the variability in previous research and the resolution of the results from this meta-analysis, it remains impossible to clarify the specific contributions within the thalamus. Brain areas within the cortical default mode network coactivate with the amygdala [27, 66] and the cerebellum [28, 60, 67, 68]. The coactivation with the amygdala involves variable groups of nuclei, including nuclei in the laterobasal, superficial, and centromedial areas [27, 66, 69, 70]. The coactivation with the cerebellum has mainly involved the CrusI/II and lobule and vermis IX of the cerebellum [19, 28, 60, 67, 68, 71], although lobules VIIIB and X have also been identified [60, 71].

We find five consistent subcortical brain regions of the default mode network. Yet, we acknowledge that additional subcortical brain regions may exist within the network that remain undetected in both our research and the broader literature, thereby limiting their representation in our meta-analytical findings. Although we performed the systematic review using only studies with volume- or combined volume- and surface-based analysis approaches, several methodological factors, as outlined in our introduction, could still account for the under-representation of subcortical involvement in the default mode network [15].

First, as we previously discussed, the quality of signals influences the detection of functional connectivity between brain regions [22]. Given that the signal-to-noise ratio is relatively low in specific regions, particularly in subcortical structures [45, 46], some cortical-subcortical connections might be overlooked in functional connectivity studies. Second, the small size and high variability of subcortical structures pose a significant challenge in aligning them for group analysis [18]. As we noted earlier, this alignment process could lead to the transformation of subcortical signals by merging them with contiguous signals - a consequence that becomes increasingly impactful when only a few voxels are allocated to these regions. These previously identified methodological challenges continue to impact our ability to thoroughly map functional connectivity between brain regions, especially within subcortical structures.

The default mode network brain regions presented here are supported by anatomical connectivity. The functional connectivity of the cortical default mode network has been considered to be bolstered by anatomical connections [72–74], including the cingulum anterior and posterior tracts, the superior and inferior longitudinal fascicles, the arcuate fasciculus, the uncinate fasciculus, the frontal orbitopolar fasciculus, and the corpus callosum [4, 18, 28, 72, 75]. Further research using diffusion tensor imaging tractography has identified tracts that connect the default mode network to the thalamus and basal forebrain [18]. The thalamus is connected to the cortical default mode network by the anterior thalamic radiations, the basal forebrain by the cingulum and fornix, and the thalamus and basal forebrain are connected to each other by the bundle of Vicq D’Azyr. Other diffusion tensor imaging tractography studies define multiple tracts connecting the cerebellum to all brain lobes, including a dentate-pontine-cerebellar tract [76]. Viral tracing techniques show connections between crus I and II and the prefrontal cortex [77], and the amygdala also has anatomical projections to the hippocampus and prefrontal cortex from its basolateral complex [78, 79].

The anatomical connections between the cortical and subcortical structures of the default mode network have led to the suggestion that the Papez circuit, an anatomical network that had been linked to emotion in 1937 [80], is part of the default mode network [18, 81, 82]. From the meta-analytical default mode network structures, the thalamus, posterior cingulate cortex, and hippocampal formation, are part of the classical Papez circuit [80]. The anatomical circuit also contains the hypothalamus and the mamillary bodies in the original proposal by Papez [80]. This circuit has progressively been separated from its hypothesized function in emotion [83] and its anatomical study has made it evolve into a more complex network that is relevant for several neurologic conditions, such as Alzheimer’s disease, and their treatments [81, 84]. Additional structures have been incorporated into the Papez circuit based on their anatomical connections, including the amygdala, which is a brain region of the meta-analytical default mode network in healthy adults and altered brain site in Alzheimer’s disease [85]. Crus I and II show anatomical connectivity with Papez circuit structures, such as thalamic nuclei, anterior cingulate cortex, and entorhinal cortex [86]. Further research should carefully examine the extent of the anatomical connections of the Papez circuit and its hypothesized functional connection to the physiology of the default mode network, with the potential of greatly impacting the ongoing development of therapeutic interventions.

The cortical default mode network as well as various subcortical regions display altered functional connectivity in Alzheimer’s disease [31, 32], but the connection between the subcortical default mode network and Alzheimer’s disease has not been elucidated yet. In the current study, we found decreased functional connectivity in a cluster in the left hippocampus, amygdala, and parahippocampal gyrus, and we empirically validated the functional connectivity of this cluster to the default mode network. This finding resonates with previous research that has found decreased functional connectivity in the default mode network in Alzheimer’s disease [35], and also with research that had found that these decreases were left-lateralized in the posterior default subnetwork [87]. Changes in the default mode network have also shown a connection to Alzheimer’s disease pathology [88]. Higher levels of amyloid-beta deposition have been found in default mode network regions [89]. The functional connectivity cluster that showed increased functional connectivity in Alzheimer’s disease was connected to a mixture of visual and ventral attention or salience networks’ areas. The anterior insula has been associated with the ventral attention and salience networks. From these often overlapping networks, the ventral attention network is right-lateralized while the salience network is bilateral [90, 91] and has shown functional connectivity increases in Alzheimer’s disease in several reports [92].

These findings also revealed a convergence between the diminished functional connectivity cluster observed in Alzheimer’s disease and the subcortical component of the default mode network. This particularly encompassed the left anterior hippocampus and the left amygdala. Prior research established a reduction in functional connectivity between the left amygdala and the bilateral precuneus when comparing Alzheimer’s disease patients exhibiting depressive symptoms to healthy controls [93]. In addition, a comprehensive evaluation of volume changes in Alzheimer’s disease indicated a significant decrease in volumes in several structures including the left amygdala, and the bilateral hippocampus [94]. A distinct correlation was detected between the presence of APOE-*ϵ*4 alleles and reduced volumes in the left amygdala and bilateral hippocampus [95]. Furthermore, there has been found an inverse relationship between tau brain accumulation and left amygdala shape [96]. The data presented herein, coupled with the cited studies, indicate that the decreased functional connectivity and reduced volumes in the left amygdala and left hippocampus might constitute a neurobiological signature of Alzheimer’s disease.

Results of the functional connectivity alteration in Alzheimer’s disease do not show alterations in cortical or subcortical default mode network brain regions outside of the hippocampal formation and amygdala, which is not in line with some previous research [31, 32, 42]. Previous research suggested that cortical regions, including the precuneus/posterior cingulate cortex and the ventromedial prefrontal cortex, are altered in their functional connectivity to other brain regions [35]. Moreover, the thalamus is associated with early pathological protein aggregations in Alzheimer’s disease [30, 97]. The absence of alterations in the thalamus and other subcortical default mode network structures could be due to a lack of shared data, small sample sizes of some studies, heterogeneous patient groups in which there are different stages of cognitive decline [98], or other methodological issues. To gain a clearer understanding of these issues, we recommend future studies involving higher-resolution data and larger sample sizes. Notwithstanding these discrepancies, the network analysis from the brain region of decreased functional connectivity yielded a functional connectivity map largely in line with the default mode network spatial pattern.

A potential limitation of this study is that the data is constrained by the availability of coordinates or brain maps in previous research. This issue may introduce a bias if certain brain structures are more commonly found in the default mode network in studies that make their coordinates or brain maps available, than in studies that do not. Coordinates are more often available than whole-brain statistical maps. This leads to lower spatial resolution in the data and meta-analysis results, making it difficult to identify the specific location of a brain region in small structures such as the thalamus. To address this issue, it is important to continue promoting open research, including the use of open repositories for neuroimaging data such as NeuroVault, openfMRI, and OpenNeuro [48, 99, 100]. It is also possible that these limitations have resulted in the exclusion of some brain regions of the default mode network. Previous research found subcortical functional brain regions of the default mode network that are not displayed in the results of the meta-analysis of the default mode network brain regions in healthy adults. For example, although most studies had shown that the cerebellar contributions to the default mode network come from the Crus I/II and lobule IX [60, 68, 71], some studies pointed to that vermis X, lobule VIIIB, or the dentate gyrus of the cerebellum, could also take part in the network [60, 71, 101]. Also, some previous studies had found functional connectivity with the basal forebrain, the caudate nucleus, the nucleus accumbens, and the ventral tegmental area [18, 19]. However, these regions may not be included in the meta-analytical default mode network map due to, for example, non-overlap of coordinates or relatively small sample sizes in some studies (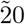 subjects for instance [18]).

This study has advanced our understanding of the subcortical structures that make up the default mode network and its alteration in Alzheimer’s disease. This study has also identified both cortical and subcortical brain regions that exhibit functional connectivity changes in Alzheimer’s disease, as well as the direction of the changes and their connection to whole-brain intrinsic functional connectivity. The results also highlight the need for further research on the connectivity of subcortical substructures to the default mode network. The participation of small subcortical regions and subparts of subcortical regions, as well as the extent of alterations in the subcortical default mode network in Alzheimer’s disease, remain unclear. Further research is needed to fully understand the role of these subcortical structures in the default mode network and how they are affected by Alzheimer’s disease. It is also important to continue promoting open research practices in order to improve the spatial resolution of meta-analytical studies and to ensure a more comprehensive and unbiased view of these issues.

## Data Availability

All brain maps derived from this study will be made available on NeuroVault upon peer-review publication.

## Abbreviations

AD,: Alzheimer’s dementia;
HC,: healthy controls;
FC,: functional connectivity

