## Supplementary material for "The Subcortical Default Mode Network and Alzheimer’s Disease: A systematic review and Activation Likelihood Estimation Meta-Analysis"

#### 1 Supplementary methods

##### 1.1 Data Extraction

Coordinates from five of the experiments (corresponding to four studies [2, 55, 34, 13]) were extracted from atlases, and one thresholded t-statistical map provided in the NeuroVault database [23], Cole's lab Github repository, the Choi's striatal parcellation in the Freesurfer wiki, and the Smith's networks at the FMRIB (Oxford) webpage. The coordinates from the five maps obtained from databases were extracted using FMRIB Software Library (FSL) [33], and AFNI tools [14, 15]. The foci of local maxima were extracted in the case of having a t-statistical map [46], and the foci of local centers of mass were extracted taking into account the distance to the cluster boundaries in the case of having an atlas. Coordinates from clusters with sizes greater than 10 voxels were included in the analysis. Coordinates originally reported in Talairach space were transformed into MNI space using the icbm2tal tool implemented by the BrainMap project in their application GingerALE [40]. The GingerALE implementation of this tool transforms coordinates in MNI space to Talairach space, and vice versa.

##### 1.2 Data set used for empirical validation and network analysis

The initial data set consisted of 184 participants, but 12 were excluded due to Quality Control issues identified within the Human Connectome Project (Quality Control issues recognized as A, B, C, and D).

Neuroimaging data were acquired with a Siemens Magnetom 7T MR Scanner and a Nova 32 32-channel Siemens receive head coil from Nova Medical. Two 16-min-long resting-state sessions were used per participant, in which one resting-state session was acquired in an anterior-to-posterior phase direction, and the other resting-state session was acquired in a posterior-to-anterior phase direction (rs-fMRI 1 and 2). Participants were instructed to fix their sight on a white cross-hair over a dark background [56]. The volumes were acquired using Gradient-Echo EPI, with a multiband factor of 5, and 85 slices per volume. The slice thickness was 1.6 mm with no gap, and the field of view (FOV) was 208 x 208 mm. The repetition time (TR) was 1000 ms, echo time (TE) was 22.2 ms, and flip angle was 45 degrees.

##### 1.3 Preprocessing of the data sets used for validation and network analysis

The downloaded HCP 7T data was already preprocessed with the HCP minimal preprocessing pipelines [22]. This included estimating transformations to reduce head motion using FSL MCFLIRT [32], applying fieldmap and gradient distortion corrections, and estimating non-linear transformations from fMRI to MNI space. The pipelines also minimize smoothing by preserving the native space resolution in the transformation to MNI, and by combining and applying all transformations in a single step using sinc interpolation. The data in MNI space were temporally filtered using a 2000 s high-pass filter, and denoised using FIX [26, 54]. The voxels in the resulting data had an isotropic resolution of 1.6 mm. CSF and WM signals were extracted and regressed using each session's `wmparc` file.

#### 2 Supplementary tables

Table 1: Search queries and the number of records found per database and repository for the default mode network meta-analysis in healthy people.

| Default mode network in healthy people | | | |
|---|---|---|---|
| Database (date) | Query # | Query | # of records |
| PubMed (05/29/2022) | 1 | (resting-state fMRI OR rsfMRI OR rs-fMRI OR task-free fMRI OR default mode network OR default network) AND (subcortical OR nucleus OR nuclei OR thalamus OR thalamic OR basal ganglia OR striatum OR striatal OR amygdala OR putamen OR pallidus OR caudate OR pons OR brainstem OR cerebellum OR hypothalamus) NOT (patient OR patients OR elders OR disorder OR dementia OR disease OR children OR childhood OR adolescence OR adolescents OR mice OR rat OR mouse OR monkey OR macaque OR rhesus OR cat) | 263 |
| Web of Science (05/29/2022) | 1 | TS=((resting-state fMRI OR rsfMRI OR rs-fMRI OR task-free fMRI OR default mode network OR default network) AND (subcortical OR nucleus OR nuclei OR thalamus OR thalamic OR basal ganglia OR striatum OR striatal OR amygdala OR putamen OR pallidus OR caudate OR pons OR brainstem OR cerebellum OR hypothalamus) NOT (patient OR patients OR elders OR disorder OR dementia OR disease OR children OR childhood OR adolescence OR adolescents OR mice OR rat OR mouse OR monkey OR macaque OR rhesus OR cat)) | 1,131 |
| Scopus (05/29/2022) | 1 | TITLE-ABS-KEY((resting-state fMRI OR rsfMRI OR rs-fMRI OR task-free fMRI OR default mode network OR default network) AND (subcortical OR nucleus OR nuclei OR thalamus OR thalamic OR basal ganglia OR striatum OR striatal OR amygdala OR putamen OR pallidus OR caudate OR pons OR brainstem OR cerebellum OR hypothalamus) AND NOT (patient OR patients OR elders OR disorder OR dementia OR disease OR children OR childhood OR adolescence OR adolescents OR mice OR rat OR mouse OR monkey OR macaque OR rhesus OR cat)) | 84 |
| NeuroVault (05/29/2022) | 1 | default network subcortical | 7 |
| | 2 | default network | 100 |
| | All databases total | | 1,585 |

Table 2: Search queries and the number of records found per database and repository for the meta-analysis of Alzheimer's disease altered brain sites

| Brain sites of functional connectivity alteration in Alzheimer's disease | | | |
|---|---|---|---|
| Database (date) | Query # | Query | # of records |
| PubMed (06/01/2022) | 1 | ((resting-state fMRI OR rsfMRI OR rs-fMRI OR task-free fMRI OR default mode network OR default network)) AND ((subcortical OR nucleus OR nuclei OR thalamus OR thalamic OR basal ganglia OR striatum OR striatal OR amygdala OR putamen OR pallidus OR caudate OR pons OR brainstem OR cerebellum OR hypothalamus)) AND ((Alzheimer's disease OR Alzheimer disease OR Alzheimer's dementia OR Alzheimer dementia dementia of the Alzheimer type OR dementia of the Alzheimer's type)) | 290 |
| WoS (06/01/2022) | 1 | TS=(((resting-state fMRI OR rsfMRI OR rs-fMRI OR task-free fMRI OR default mode network OR default network)) AND ((subcortical OR nucleus OR nuclei OR thalamus OR thalamic OR basal ganglia OR striatum OR striatal OR amygdala OR putamen OR pallidum OR caudate OR pons OR brainstem OR cerebellum OR hypothalamus)) AND ((Alzheimer's disease OR Alzheimer disease OR Alzheimer's dementia OR Alzheimer dementia dementia of the Alzheimer type OR dementia of the Alzheimer's type))) | 357 |
| Scopus (06/01/2022) | 1 | TITLE-ABS-KEY ( ( ( resting-state AND fmri OR rsfmri OR rs-fmri OR task-free AND fmri OR default AND mode AND network OR default AND network ) ) AND ( ( subcortical OR nucleus OR nuclei OR thalamus OR thalamic OR basal AND ganglia OR striatum OR striatal OR amygdala OR putamen OR pallidus OR caudate OR pons OR brainstem OR cerebellum OR hypothalamus ) ) AND ( ( alzheimer's AND disease OR alzheimer AND disease OR alzheimer's AND dementia OR alzheimer AND dementia AND dementia AND of AND the AND alzheimer AND type OR dementia AND of AND the AND alzheimer's AND type ) ) ) | 0 |
| NeuroVault (06/01/2022) | 1 | default Alzheimer | 4 |
| | All databases total | | 651 |

Table 3: Included studies and experiments for meta-analysis of the default mode network in healthy adults.

| Study | Exp # | Sample | | | DMN extraction | | |
|---|---|---|---|---|---|---|---|
| | | N | Age (mean (SD) or range) | Gender (male) | Network extraction technique | Initial coordinate space | subcortical foci |
| Alves et al. (2019)[2] | 1 | 20 | 29 (6) | 45% | seed-based FC | MNI | thalamus, basal forebrain, mammillary bodies, midbrain, caudate nucleus |
| | 2 | 20 | 29 (6) | 45% | seed-based FC | MNI | |
| Smith et al. (2009)[55] | 1 | 36 | range: 20 - 35 | 58% | ICA | MNI | none |
| Choi et al. (2012)[13] | 1 | 500 | range: 18 - 35 | 43% | voxel-wise FC | MNI | striatum |
| Ji et al. (2020)[34] | 1 | 337 | range: 22 - 35 | N/A | voxel-wise FC | MNI | |
| Bzdok et al. (2013)[10] | 1-2 | 139 | 42.30 | 60% | seed-based FC | MNI | amygdala, nucleus accumbens, and cerebellum |
| Buckner et al. (2011)[9] | 1 | 500 | range: 18 - 35 | 43% | voxel-wise FC | MNI | cerebellum |
| Bernard et al. (2014)[7] | 1-4 | 31 | 22.76 (2.95) | 48% | seed-based FC | MNI | cerebellum (dentate gyrus), brainstem, putamen, caudate |
| Chase et al. (2020)[12] | 1-4 | 196 | 39.80 (15.1) | 61% | seed-based FC | MNI | amygdala, caudate, and thalamus |
| Fang et al. (2020)[21] | 1-2 | 35 | 23.69 (3.57) | 43% | seed-based FC | MNI | pons, cerebellum, and caudate |
| Li et al. (2013)[43] | 1-2 | 20 | range: 19 - 28 | 40% | seed-based FC | MNI | caudate, cerebellum (crusI/II, lobule IX) |
| Grigg and Grady (2010)[27] | 1 | 20 | 23.70 (3) | 50% | seed-based FC | MNI | caudate, thalamus, cerebellum |
| Jung et al. (2014)[36] | 1-4 | 59 | 25.81 (6.45) | 56% | seed-based and k-means | MNI | caudate, putamen |
| Blessing et al. (2016)[8] | 1 | 50 | 24.30 (4.10) | 54% | masked ICA and DR | MNI | nucleus acumbens, amygdala, cerebellum (crusI/II, lobule and vermis IX, lobule VIIIB and X), thalamus, ventral tegmental area |
| Kawabata et al. (2022)[38] | 1 | 101 | range: 20 - 49 | 37% | FCOR | MNI | cerebellum (crus I/II, lobule IX, vermis IX, X) |
| Zhou et al. (2011)[67] | 1 | 25 | 29.44 (8.47) | 40% | PCA-ICA | Talairach | none |
| Gultepe et al. (2013)[28] | 1 | 17 | range: 21 - 39 | 47% | seed-based FC | MNI | cerebellum (tonsil and crus) |
| Fan et al. (2021)[20] | 1-6 | 100 | range: 22 - 35 | N/A | dynamic FC degree, ICA | "Talairach MNI" | none |
| Bär et al. (2016)[5] | 1-4 | 154 | 28.80 (9.60) | 47% | seed-based FC | MNI | cerebellum (uvula, declive), ventral tegmental area, nucleus raphes dorsalis, substantia nigra |
| | 5 | 154 | 28.80 (9.60) | 47% | graph theory | MNI | cerebellum (uvula, declive), ventral tegmental area, nucleus raphes dorsalis, and substantia nigra |
| | 6-7 | 77 | ∼28.8(9.60) | ∼47% | graph theory | MNI | cerebellum (uvula, declive), ventral tegmental area, nucleus raphes dorsalis, and substantia nigra |
| Lee and Xue (2018) [41] | 1-3 | 36 | 27.40 range: 19 - 42 | 50% | seed-based FC | Talairach | cerebellum, thalamus, pons |
| Kark et al. (2021)[37] | 1 | 121 | range: 22 - 35 | 40% | seed-based FC | MNI | thalamus |
| Tsai et al. (2014)[59] | 1 | 22 | 23.80 (4.20) | 100% | seed-based FC | MNI | thalamus |
| Wang et al. (2016)[61] | 1 | 24 | 20.63 (3.20) | 46% | Full correlation matrix | MNI | none |
| Bernard et al. (2012) [6] | 1 | 39 | 22.76 (2.95) | 56% | seed-based FC | MNI | thalamus |
| Andrade et al. (2011) [3] | 1 | 25 | 24.70 (3.20) | 52% | seed-based FC | MNI | none |
| de la Cruz et al. (2019) [17] | 1 | 84 | 31.58 (10.76) | 49% | seed-based FC | MNI | amygdala, cerebellum |
| Jarrahi et al. (2016) [31] | 1-4 | 15 | range: 21 - 48 | 0% | ICA | MNI | none |

Abbreviations: DMN, default mode network; DR, dual regression; FC, functional connectivity; FCOR, functional connectivity overlap ratio; ICA, Independent Component Analysis; MNI, Montreal Neurological Initiative; PCA, principal component analysis.

Table 4: Included studies and experiments for meta-analysis of altered brain sites in Alzheimer's disease, in which AD <HC.

| Study | Exp # | Sample | | | | | | AD diagnosis | Data analysis |
|---|---|---|---|---|---|---|---|---|---|
| | | HC | | | AD | | | | analysis |
| | | N | Age (mean (SD) or range) | Gender (male) | N | Age (mean (SD) or range) | Gender (male) | | |
| Balachandar et al. 2013[4] | 1 | 15 | 64,4 (8,9) | 60% | 15 | 67,33 (6,6) | 60% | mild AD[48] | ICA + DR |
| Qi et al., 2019[52] | 1 | 26 | 71,3 (6,8) | 54% | 20 | 73,1 (6,7) | 45% | probable AD (NINCDS-ADRDA) | seed-based FC |
| Zheng et al., 2017[65] | 1 | 38 | 68.39 (7.78) | 34% | 32 | 71.25 (8.63) | 44% | possible/probable AD[18, 19] | seed-based FC |
| Multani et al., 2019[49] | 1 | 10 | 62.5 (5.5) | 40% | 18 | 70.56 (10.4) | 39% | AD[48] | seed-based FC |
| Chabran et al., 2020[11] | 1 | 22 | 66.5 (7.8) | 50% | 58 | 73.7 (8.3) | 37% | AD[18] | ROI-to-ROI FC |
| Zhou et al., 2021[68] | 1 | 42 | 69.86 (6.66) | 43% | 46 | 73.17 (7.09) | 41% | probable AD[48] | seed-based FC |
| Yao et al., 2013[63] | 1-2 | 27 | 69.2 (6.5) | 59% | 35 | 72.4 (8.5) | 34% | probable AD[57] | seed-based FC |
| Zhang et al., 2009[64] | 1 | 16 | 71.3 (4.9) | 44% | 16 | 71.6 (5.5) | 38% | possible/probable AD (NINCDS-ADRDA) | seed-based FC |
| Tang et al., 2021[58] | 1-4 | 20 | 62.6 (6.95) | 55% | 27 | 64.81 (8.24) | 41% | AD[48] | seed-based FC |
| Grajski et al., 2019[25] | 1-2 | 32 | 74.2 (5.5) | 44% | 34 | 73.4 (7.2) | 62% | ADNI AD | ROI-to-ROI FC |
| Wang et al., 2016[62] | 1 | 38 | 68.39 (7.78) | 34% | 32 | 71.25 (8.63) | 44% | possible/probable AD | seed-based FC |
| Altunkaya et al., 2020[1] | 1 | 23 | 65.7 (7.42) | 52% | 34 | 77.8 (6.51) | 50% | AD[48] | constrained group ICA |
| Kim et al., 2016[39] | 1 | 65 | 69.9 (3.9) | 34% | 37 | 66.8 (8.7) | 38% | probable AD[47] | group ICA |
| Zhou et al., 2010[66] | 1-2 | 12 | 62.0 (8.9) | 42% | 12 | 63.3 (7.7) | 42% | probable AD[48] | ICA |
| Ji et al., 2021[35] | 1 | 12 | 64.83 (6.94) | 58% | 12 | 67.58 (9.05) | 58% | probable AD (NINCDS-ADRDA) | group ICA |
| Gour et al., 2014[24] | 1-8 | 14 | 72.8 (3) | 29% | 14 | 75.1 (2.9) | 43% | probable AD[48] | seed-based FC |
| Dai et al., 2015[16] | 1 | 38 | 68.39 (7.78) | 34% | 32 | 71.25 (8.63) | 44% | possible/probable AD [48, 18, 19] | nodal FC strenght |
| Wang et al., 2015[60] | 1 | 16 | 69.25 (7.83) | 44% | 16 | 71.56 (5.93) | 44% | possible/probable AD[47] | VMHC |
| Mascali et al., 2015[45] | 1 | 10 | 66.0 (9.6) | 70% | 10 | 72.3 (8.3) | 40% | possible/probable AD[47] | coupling between FC and ALFF |
| Herdick et al., 2020[30] | 1-2 | 174 | 68.9 (5.2) | 41% | 51 | 73.0 (6.6) | 43% | probable AD[48] | seed-based FC |
| Qiao et al., 2018[53] | 1 | 34 | 65.55 (8.98) | 62% | 34 | 68.64 (9.85) | 50% | AD[48] | 3LHPM-ICA |
| Lee et al., 2020[42] | 1 | 25 | 71.56 (5.98) | 52% | 57 | 75.11 (8.38) | 33% | AD[47] | seed-based FC |
| Ortner et al., 2016[50] | 1 | 33 | 56.15 (9.31) | 36% | 38 | 62.86 (8.90) | 50 | probable AD[48] | seed-based FC |
| Peraza et al., 2016[51] | 1 | 17 | 76.75 (5.93) | 81% | 24 | 75.39 (8.6) | 83% | AD (NINCDS-ADRDA) | ReHo |
| Ma et al., 2022[44] | 1 | 33 | 64.94 (9.39) | 45% | 34 | 65.88 (8.26) | 41% | probable AD[48] | node degree |
| Hafkemeijer et al., 2015[29] | 1 | 29 | 62.8 (5.1) | 59% | 31 | 65.3 (7.0) | 76% | probable AD[48] | network-to-ROI and ROI-to-ROI FC |

Abbreviations: 3LHPM, three-level hierarchical partner matching; AD, Alzheimer's disease; ALFF, amplitude of low-frequency fluctuations; FC, functional connectivity; GLM, general linear model; ICA, independent component analysis; ReHo, regional homogeneity; ROI, region of interest; VMHC, voxel-mirrored homotopic connectivity.

Table 5: Included studies and experiments for meta-analysis of altered brain sites in Alzheimer's disease, in which AD <HC.

| Study | Exp # | Sample | | | | | | | Data analysis |
|---|---|---|---|---|---|---|---|---|---|
| | | HC | | | AD | | | AD diagnosis | analysis |
| | | N | Age (mean (SD) or range) | Gender (male) | N | Age (mean (SD) or range) | Gender (male) | | |
| Balachandar et al. 2013[4] | 1 | 15 | 64,4 (8,9) | 60% | 15 | 67,33 (6,6) | 60% | mild AD[48] | ICA + DR |
| Qi et al., 2019[52] | 1 | 26 | 71,3 (6,8) | 54% | 20 | 73,1 (6,7) | 45% | probable AD (NINCDS-ADRDA) | seed-based FC |
| Zheng et al., 2017[65] | 1 | 38 | 68.39 (7.78) | 34% | 32 | 71.25 (8.63) | 44% | possible/probable AD[18, 19] | seed-based FC |
| Multani et al., 2019[49] | 1 | 10 | 62.5 (5.5) | 40% | 18 | 70.56 (10.4) | 39% | AD[48] | seed-based FC |
| Chabran et al., 2020[11] | 1 | 22 | 66.5 (7.8) | 50% | 58 | 73.7 (8.3) | 37% | AD[18] | ROI-to-ROI FC |
| Zhou et al., 2021[68] | 1 | 42 | 69.86 (6.66) | 43% | 46 | 73.17 (7.09) | 41% | probable AD[48] | seed-based FC |
| Yao et al., 2013[63] | 1-2 | 27 | 69.2 (6.5) | 59% | 35 | 72.4 (8.5) | 34% | probable AD[57] | seed-based FC |
| Zhang et al., 2009[64] | 1 | 16 | 71.3 (4.9) | 44% | 16 | 71.6 (5.5) | 38% | possible/probable AD (NINCDS-ADRDA) | seed-based FC |
| Tang et al., 2021[58] | 1-4 | 20 | 62.6 (6.95) | 55% | 27 | 64.81 (8.24) | 41% | AD[48] | seed-based FC |
| Grajski et al., 2019[25] | 1-2 | 32 | 74.2 (5.5) | 44% | 34 | 73.4 (7.2) | 62% | ADNI AD | ROI-to-ROI FC |
| Wang et al., 2016[62] | 1 | 38 | 68.39 (7.78) | 34% | 32 | 71.25 (8.63) | 44% | possible/probable AD | seed-based FC |
| Altunkaya et al., 2020[1] | 1 | 23 | 65.7 (7.42) | 52% | 34 | 77.8 (6.51) | 50% | AD[48] | constrained group ICA |
| Kim et al., 2016[39] | 1 | 65 | 69.9 (3.9) | 34% | 37 | 66.8 (8.7) | 38% | probable AD[47] | group ICA |
| Zhou et al., 2010[66] | 1-2 | 12 | 62.0 (8.9) | 42% | 12 | 63.3 (7.7) | 42% | probable AD[48] | ICA |
| Ji et al., 2021[35] | 1 | 12 | 64.83 (6.94) | 58% | 12 | 67.58 (9.05) | 58% | probable AD (NINCDS-ADRDA) | group ICA |
| Gour et al., 2014[24] | 1-8 | 14 | 72.8 (3) | 29% | 14 | 75.1 (2.9) | 43% | probable AD[48] | seed-based FC |
| Dai et al., 2015[16] | 1 | 38 | 68.39 (7.78) | 34% | 32 | 71.25 (8.63) | 44% | possible/probable AD [48, 18, 19] | nodal FC strenght |
| Wang et al., 2015[60] | 1 | 16 | 69.25 (7.83) | 44% | 16 | 71.56 (5.93) | 44% | possible/probable AD[47] | VMHC |
| Mascali et al., 2015[45] | 1 | 10 | 66.0 (9.6) | 70% | 10 | 72.3 (8.3) | 40% | possible/probable AD[47] | coupling between FC and ALFF |
| Herdick et al., 2020[30] | 1-2 | 174 | 68.9 (5.2) | 41% | 51 | 73.0 (6.6) | 43% | probable AD[48] | seed-based FC |
| Qiao et al., 2018[53] | 1 | 34 | 65.55 (8.98) | 62% | 34 | 68.64 (9.85) | 50% | AD[48] | 3LHPM-ICA |
| Lee et al., 2020[42] | 1 | 25 | 71.56 (5.98) | 52% | 57 | 75.11 (8.38) | 33% | AD[47] | seed-based FC |
| Ortner et al., 2016[50] | 1 | 33 | 56.15 (9.31) | 36% | 38 | 62.86 (8.90) | 50 | probable AD[48] | seed-based FC |
| Peraza et al., 2016[51] | 1 | 17 | 76.75 (5.93) | 81% | 24 | 75.39 (8.6) | 83% | AD (NINCDS-ADRDA) | ReHo |
| Ma et al., 2022[44] | 1 | 33 | 64.94 (9.39) | 45% | 34 | 65.88 (8.26) | 41% | probable AD[48] | node degree |
| Hafkemeijer et al., 2015[29] | 1 | 29 | 62.8 (5.1) | 59% | 31 | 65.3 (7.0) | 76% | probable AD[48] | network-to-ROI and ROI-to-ROI FC |

Abbreviations: 3LHPM, three-level hierarchical partner matching; AD, Alzheimer's disease; ALFF, amplitude of low-frequency fluctuations; FC, functional connectivity; GLM, general linear model; ICA, independent component analysis; ReHo, regional homogeneity; ROI, region of interest; VMHC, voxel-mirrored homotopic connectivity.

Table 6: Coordinates for the ten clusters in the functional connectivity map from the default mode network meta-analytical mask, at threshold t > 7, in the HCP 7T dataset.

| Number of voxels | Peak t statistic | Peak X (voxel) | Peak Y (voxel) | Peak Z (voxel) | COG X (voxel) | COG Y (voxel) | COG Z (voxel) |
|---|---|---|---|---|---|---|---|
| 19326 | 24.5 | 57 | 70 | 50 | 58.5 | 103 | 58.5 |
| 12690 | 37.4 | 57 | 43 | 59 | 56.1 | 48.9 | 57.5 |
| 4357 | 20.1 | 95 | 78 | 32 | 91 | 68.7 | 35.2 |
| 3307 | 31 | 88 | 35 | 67 | 84.7 | 38.8 | 65.4 |
| 3187 | 27.2 | 36 | 31 | 24 | 37.4 | 31.4 | 23.2 |
| 3126 | 18 | 20 | 73 | 37 | 20.7 | 73.2 | 33.6 |
| 2700 | 31.8 | 23 | 40 | 64 | 25.2 | 41.5 | 64.7 |
| 2327 | 21.4 | 78 | 32 | 23 | 74.5 | 30.7 | 22.6 |
| 1157 | 31.5 | 59 | 44 | 16 | 56 | 45.5 | 17.4 |
| 932 | 17.4 | 39 | 89 | 34 | 31.8 | 99.1 | 36.6 |
| 338 | 12.4 | 35 | 62 | 78 | 33.3 | 64 | 80.2 |
| 229 | 11 | 25 | 71 | 69 | 22.1 | 73.8 | 64.2 |
| 213 | 10.4 | 77 | 60 | 80 | 78.3 | 62.2 | 80.8 |
| 206 | 10.7 | 91 | 73 | 66 | 89.4 | 72.2 | 64.6 |
| 150 | 12 | 60 | 63 | 81 | 58.3 | 62.1 | 83.5 |
| 146 | 13.3 | 54 | 62 | 82 | 53.5 | 62.6 | 83.9 |
| 118 | 11.2 | 48 | 58 | 87 | 48.5 | 59 | 88.2 |
| 116 | 19.6 | 56 | 65 | 32 | 55.9 | 63.6 | 32.5 |
| 109 | 11.1 | 64 | 58 | 85 | 63 | 58.6 | 87.8 |
| 24 | 8.72 | 20 | 96 | 53 | 20.5 | 95.5 | 53.3 |
| 24 | 8.77 | 81 | 71 | 29 | 80.8 | 70.2 | 28.7 |
| 17 | 11.2 | 54 | 75 | 39 | 54.3 | 74.2 | 38.5 |
| 16 | 9.67 | 35 | 63 | 55 | 34.5 | 64.4 | 55.3 |
| 14 | 9.67 | 79 | 88 | 27 | 78.9 | 88.1 | 27.1 |
| 14 | 11.6 | 64 | 60 | 31 | 64.3 | 60.1 | 30.7 |
| 11 | 8.57 | 78 | 60 | 57 | 77.6 | 61.3 | 55.5 |

Table 7: Coordinates of the functional connectivity map to the default mode network meta-analytical cluster 1 (PCC, precuneus), at threshold t > 7, in the HCP 7T dataset

| Number of voxels | Peak t statistic | Peak X (voxel) | Peak Y (voxel) | Peak Z (voxel) | COG X (voxel) | COG Y (voxel) | COG Z (voxel) |
|---|---|---|---|---|---|---|---|
| 12381 | 23 | 56 | 116 | 42 | 58.3 | 106 | 57.8 |
| 10837 | 50.5 | 57 | 43 | 59 | 57.6 | 46.9 | 59.3 |
| 3239 | 19.2 | 94 | 72 | 39 | 91.6 | 69.4 | 35.2 |
| 2809 | 26.8 | 88 | 35 | 67 | 84 | 37.9 | 65.5 |
| 1918 | 25 | 23 | 40 | 64 | 25.7 | 40.4 | 64.2 |
| 1840 | 19 | 32 | 33 | 22 | 37.2 | 31.1 | 22.5 |
| 1795 | 17.4 | 16 | 78 | 34 | 20.7 | 76.3 | 32.5 |
| 1303 | 21.9 | 41 | 67 | 35 | 40.3 | 64.7 | 34.3 |
| 1047 | 17.9 | 81 | 33 | 22 | 73.9 | 29.7 | 22.2 |
| 963 | 19.3 | 54 | 44 | 14 | 55.5 | 45.7 | 17.2 |
| 399 | 13.6 | 80 | 99 | 36 | 79.5 | 96.1 | 35.9 |
| 370 | 16.3 | 33 | 101 | 38 | 33.4 | 98.7 | 36.5 |
| 255 | 10.8 | 35 | 62 | 78 | 33.2 | 63.7 | 80.5 |
| 201 | 12.6 | 61 | 61 | 51 | 60.2 | 63.9 | 50.3 |
| 136 | 10.7 | 25 | 71 | 69 | 22.1 | 73.3 | 66 |
| 126 | 09.02 | 79 | 62 | 85 | 78.8 | 62.3 | 80.8 |
| 125 | 13.2 | 54 | 61 | 82 | 53.6 | 62.4 | 83.6 |
| 98 | 11 | 60 | 63 | 81 | 58.4 | 61.8 | 83.6 |
| 95 | 9.68 | 85 | 93 | 59 | 87.7 | 93.4 | 55.3 |
| 90 | 9.6 | 92 | 73 | 67 | 89.8 | 72.2 | 64.9 |
| 82 | 17 | 56 | 64 | 32 | 56 | 63.6 | 32.2 |
| 73 | 10.1 | 48 | 58 | 86 | 48.5 | 58.5 | 87.9 |
| 73 | 10.6 | 62 | 57 | 86 | 63 | 58.7 | 87.9 |
| 49 | 13.1 | 50 | 61 | 51 | 50.3 | 61.2 | 50.4 |
| 22 | 9.41 | 82 | 33 | 16 | 81.1 | 33.5 | 15.3 |
| 17 | 9.28 | 35 | 86 | 28 | 34.1 | 87.2 | 27.8 |
| 14 | 9.34 | 66 | 35 | 26 | 65.4 | 34.7 | 26.6 |

Table 8: Coordinates of the functional connectivity map to the default mode network meta-analytical cluster 2 (MTG, STG), at threshold t > 7, in the HCP 7T dataset.

| Number of voxels | Peak t statistic | Peak X (voxel) | Peak Y (voxel) | Peak Z (voxel) | COG X (voxel) | COG Y (voxel) | COG Z (voxel) |
|---|---|---|---|---|---|---|---|
| 14762 | 21.3 | 70 | 95 | 71 | 61.1 | 104 | 60.2 |
| 8182 | 29.9 | 56 | 37 | 66 | 56.3 | 45.5 | 62.7 |
| 4261 | 18.8 | 95 | 57 | 45 | 91.4 | 67.3 | 35.5 |
| 3447 | 37.1 | 88 | 36 | 66 | 84.5 | 39 | 65.9 |
| 2954 | 19.1 | 32 | 34 | 22 | 37.5 | 31.4 | 23.1 |
| 2311 | 27.6 | 23 | 40 | 64 | 25.5 | 41.1 | 65.2 |
| 2306 | 16 | 17 | 72 | 39 | 19.8 | 72.9 | 33.9 |
| 1402 | 18.6 | 79 | 33 | 22 | 74.9 | 30.2 | 22.2 |
| 1071 | 21.3 | 72 | 67 | 35 | 71.4 | 63.1 | 34.4 |
| 900 | 18.8 | 53 | 46 | 17 | 55.5 | 45.5 | 17.3 |
| 855 | 19.6 | 39 | 68 | 34 | 40.1 | 65.2 | 33.9 |
| 553 | 13.2 | 33 | 102 | 38 | 30.8 | 101 | 36.6 |
| 99 | 9.81 | 24 | 72 | 66 | 22.6 | 73.9 | 64.1 |
| 76 | 10 | 61 | 86 | 53 | 63.5 | 85.3 | 54.5 |
| 75 | 12.5 | 61 | 61 | 51 | 61.4 | 60.3 | 49.5 |
| 75 | 10.6 | 51 | 91 | 48 | 50 | 87.8 | 52.6 |
| 61 | 11 | 54 | 62 | 83 | 53.7 | 62 | 83.4 |
| 56 | 9.6 | 91 | 73 | 66 | 89.5 | 72.3 | 65.5 |
| 56 | 9.22 | 38 | 116 | 47 | 37.8 | 114 | 47.5 |
| 53 | 10.9 | 55 | 89 | 41 | 56 | 88.1 | 41 |
| 37 | 12.8 | 51 | 62 | 51 | 50.3 | 60.8 | 50.3 |
| 35 | 8.89 | 35 | 62 | 78 | 34.4 | 62 | 80.9 |
| 35 | 13.7 | 56 | 65 | 32 | 56.3 | 65.2 | 31.9 |
| 28 | 10.5 | 66 | 35 | 26 | 66.2 | 34.2 | 26.9 |
| 15 | 8.82 | 35 | 86 | 28 | 34.6 | 86.4 | 27.5 |
| 15 | 8.65 | 58 | 80 | 45 | 58 | 81.2 | 45.5 |
| 14 | 8.59 | 77 | 40 | 26 | 76.7 | 38.7 | 26.5 |
| 13 | 8.6 | 60 | 63 | 81 | 58.5 | 61.9 | 82.4 |
| 13 | 7.95 | 57 | 69 | 51 | 57.9 | 68.3 | 51.5 |
| 11 | 7.88 | 29 | 68 | 79 | 29.5 | 68 | 78.7 |

Abbreviations: COG, Center Of Gravity.

Table 9: Coordinates of the functional connectivity map to the default mode network meta-analytical cluster 3 (ACC, MFG), at threshold t > 7, in the HCP 7T dataset.

| Number of voxels | Peak t statistic | Peak X (voxel) | Peak Y (voxel) | Peak Z (voxel) | COG X (voxel) | COG Y (voxel) | COG Z (voxel) |
|---|---|---|---|---|---|---|---|
| 8653 | 19.4 | 54 | 51 | 57 | 55.7 | 46.1 | 62 |
| 6643 | 47.1 | 50 | 104 | 42 | 55.3 | 107 | 48.8 |
| 1524 | 15.7 | 25 | 37 | 69 | 25.4 | 40.3 | 65.3 |
| 1173 | 14.1 | 41 | 98 | 75 | 40.3 | 96.5 | 72.9 |
| 915 | 13.8 | 79 | 30 | 71 | 83.6 | 35.6 | 67 |
| 551 | 13.3 | 70 | 96 | 69 | 70 | 95.7 | 73 |
| 455 | 13.1 | 71 | 66 | 36 | 71.1 | 59.5 | 35.5 |
| 398 | 14.4 | 40 | 67 | 35 | 40.5 | 61.1 | 35.6 |
| 318 | 11.9 | 63 | 49 | 17 | 57 | 46.6 | 16.2 |
| 306 | 10.7 | 83 | 35 | 22 | 83.7 | 37.5 | 20.8 |
| 247 | 10.1 | 18 | 75 | 37 | 18.3 | 73.8 | 36.3 |
| 123 | 10.4 | 49 | 62 | 52 | 53 | 65.6 | 51.3 |
| 114 | 10.6 | 63 | 26 | 19 | 64 | 25.4 | 20.2 |
| 109 | 12.2 | 39 | 90 | 35 | 36.2 | 89.5 | 36.6 |
| 61 | 8.68 | 92 | 77 | 36 | 92.1 | 75.4 | 35.6 |
| 58 | 10.9 | 77 | 88 | 37 | 76.2 | 87.8 | 36.2 |
| 52 | 8.86 | 34 | 102 | 40 | 34.6 | 101 | 37.7 |
| 50 | 14.8 | 56 | 66 | 33 | 55.9 | 65.9 | 32.9 |
| 44 | 9.36 | 26 | 35 | 22 | 26.7 | 36.3 | 20.9 |
| 31 | 8.35 | 14 | 64 | 40 | 14.7 | 63.3 | 40.8 |
| 26 | 9.76 | 30 | 43 | 18 | 28.9 | 42 | 18.7 |
| 21 | 9.41 | 82 | 34 | 14 | 82.2 | 35.2 | 14.8 |
| 20 | 9.38 | 48 | 25 | 21 | 48.2 | 25.3 | 21 |
| 16 | 10.3 | 54 | 62 | 45 | 53.7 | 63.7 | 44.8 |
| 16 | 8.7 | 69 | 28 | 30 | 72.7 | 27.4 | 28.9 |
| 15 | 9.67 | 58 | 62 | 45 | 58.1 | 63.8 | 44.7 |
| 14 | 9.58 | 76 | 40 | 26 | 77.3 | 40.3 | 25.9 |
| 11 | 8.51 | 16 | 76 | 30 | 16.5 | 76.3 | 29.9 |

Table 10: Coordinates of the functional connectivity map to the default mode network meta-analytical cluster 4 (PHG, amygdala, hippocampus), at threshold $t > 7$, in the HCP 7T dataset.

| Number of voxels | Peak t statistic | Peak X (voxel) | Peak Y (voxel) | Peak Z (voxel) | COG X (voxel) | COG Y (voxel) | COG Z (voxel) |
|---|---|---|---|---|---|---|---|
| 10872 | 17.7 | 31 | 66 | 74 | 49.8 | 66.5 | 71.6 |
| 10790 | 66.4 | 39 | 69 | 33 | 55.1 | 51.6 | 50.3 |
| 5729 | 18.6 | 70 | 94 | 70 | 57.2 | 106 | 60 |
| 2366 | 16.5 | 93 | 73 | 39 | 92.2 | 70.6 | 39.5 |
| 2323 | 20 | 85 | 34 | 64 | 83.1 | 35.6 | 64.2 |
| 1855 | 17.9 | 21 | 39 | 62 | 26.3 | 38.7 | 62.5 |
| 542 | 13.3 | 34 | 66 | 56 | 30.9 | 66.3 | 55.3 |
| 469 | 13.3 | 78 | 59 | 57 | 81.3 | 63.4 | 55.1 |
| 290 | 12.4 | 53 | 47 | 17 | 55.1 | 46.2 | 16 |
| 200 | 17.9 | 33 | 101 | 38 | 33.2 | 101 | 37.4 |
| 147 | 14.4 | 59 | 81 | 45 | 56 | 85.1 | 42.8 |
| 136 | 11.2 | 33 | 56 | 29 | 33 | 53.1 | 30.6 |
| 133 | 9.17 | 97 | 51 | 40 | 94.1 | 51.2 | 38.8 |
| 102 | 13.4 | 80 | 99 | 37 | 79.5 | 99.1 | 36.8 |
| 98 | 9.79 | 81 | 90 | 62 | 82.5 | 90.6 | 61.2 |
| 86 | 11.2 | 64 | 25 | 20 | 63.7 | 25.4 | 20.2 |
| 80 | 11.6 | 48 | 26 | 20 | 47.7 | 25.6 | 20.7 |
| 64 | 10.7 | 78 | 53 | 30 | 78.8 | 51 | 30.6 |
| 59 | 11 | 83 | 35 | 22 | 81.6 | 34 | 21.8 |
| 50 | 9.88 | 48 | 44 | 42 | 48.1 | 42.9 | 42.5 |
| 50 | 10.5 | 31 | 35 | 22 | 30.6 | 34.6 | 21.7 |
| 44 | 11.1 | 81 | 71 | 29 | 80.9 | 69.6 | 29.1 |
| 42 | 12.8 | 59 | 80 | 63 | 58.4 | 80.8 | 62.8 |
| 40 | 11.2 | 58 | 99 | 51 | 58.1 | 98.4 | 51.4 |
| 32 | 10.9 | 53 | 100 | 50 | 52.7 | 98.1 | 52.1 |
| 29 | 10.7 | 35 | 86 | 27 | 33.8 | 86.5 | 28.3 |
| 29 | 12 | 53 | 80 | 63 | 52.6 | 81.8 | 62.8 |
| 27 | 8.92 | 67 | 40 | 42 | 66.1 | 40.8 | 41.8 |
| 17 | 8.54 | 16 | 55 | 39 | 17.2 | 53.8 | 39.3 |
| 17 | 9.39 | 28 | 47 | 36 | 27.4 | 47.4 | 35.4 |
| 14 | 9.3 | 74 | 79 | 36 | 74.3 | 79.2 | 35.6 |
| 11 | 7.86 | 43 | 31 | 41 | 42.2 | 30.6 | 39.6 |

Table 11: Coordinates of the functional connectivity map to the default mode network meta-analytical cluster 5 (MTG R), at threshold t > 7, in the HCP 7T dataset.

| Number of voxels | Peak t statistic | Peak X (voxel) | Peak Y (voxel) | Peak Z (voxel) | COG X (voxel) | COG Y (voxel) | COG Z (voxel) |
|---|---|---|---|---|---|---|---|
| Number of voxels | Peak t statistic | Peak X (voxel) | Peak Y (voxel) | Peak Z (voxel) | COG X (voxel) | COG Y (voxel) | COG Z (voxel) |
| 16989 | 23.1 | 42 | 97 | 75 | 53.8 | 104 | 60.1 |
| 8816 | 29.9 | 56 | 40 | 66 | 55.8 | 45.6 | 63.3 |
| 3546 | 17 | 95 | 57 | 45 | 91.9 | 67.2 | 35.4 |
| 3340 | 39.5 | 24 | 40 | 66 | 25.4 | 42.4 | 65.9 |
| 3272 | 27.6 | 89 | 38 | 67 | 84.5 | 39.3 | 66.5 |
| 3081 | 19.8 | 81 | 35 | 22 | 74.6 | 31.5 | 22.8 |
| 3071 | 16.9 | 15 | 61 | 43 | 19.6 | 70.2 | 34.8 |
| 2624 | 16.5 | 31 | 35 | 22 | 37 | 31.7 | 23.2 |
| 996 | 19.2 | 59 | 44 | 13 | 56.3 | 45.5 | 17.3 |
| 776 | 17.8 | 39 | 66 | 36 | 40 | 64.2 | 34.4 |
| 776 | 17.8 | 72 | 67 | 35 | 71.3 | 62.4 | 34.3 |
| 166 | 13.7 | 48 | 84 | 56 | 48.5 | 85.8 | 54.4 |
| 112 | 11.4 | 61 | 86 | 53 | 63.1 | 85.6 | 54.4 |
| 110 | 10.5 | 91 | 92 | 56 | 89.2 | 92.5 | 52.8 |
| 99 | 15.5 | 50 | 61 | 51 | 50.4 | 61.4 | 50.3 |
| 92 | 11.7 | 75 | 88 | 33 | 75.6 | 90.4 | 34.1 |
| 74 | 8.96 | 20 | 94 | 56 | 21.1 | 94 | 53.6 |
| 70 | 13 | 60 | 60 | 50 | 61.2 | 60.4 | 49.8 |
| 59 | 15.3 | 56 | 65 | 32 | 55.7 | 65.8 | 32.4 |
| 26 | 9.3 | 55 | 89 | 41 | 56.2 | 88 | 41 |
| 19 | 10.1 | 54 | 70 | 52 | 53.6 | 69.9 | 51.7 |
| 14 | 9.13 | 58 | 70 | 51 | 58.1 | 68.6 | 51.7 |

Table 12: Coordinates of the functional connectivity map to the default mode network meta-analytical cluster 6 (PHG left), at threshold t > 7, in the HCP 7T dataset.

| Number of voxels | Peak t statistic | Peak X (voxel) | Peak Y (voxel) | Peak Z (voxel) | COG X (voxel) | COG Y (voxel) | COG Z (voxel) |
|---|---|---|---|---|---|---|---|
| 9493 | 79.2 | 74 | 62 | 37 | 57.2 | 49.8 | 53.5 |
| 4472 | 16.5 | 68 | 98 | 69 | 60.5 | 109 | 55.6 |
| 2155 | 18.4 | 89 | 36 | 67 | 83.6 | 36.9 | 65.6 |
| 1287 | 15.9 | 23 | 38 | 66 | 26 | 39.3 | 64.4 |
| 1112 | 12.3 | 31 | 66 | 74 | 30.9 | 67.5 | 75.6 |
| 1040 | 13.4 | 92 | 74 | 38 | 92.3 | 71.6 | 36.8 |
| 635 | 13.7 | 43 | 96 | 72 | 41.5 | 97.1 | 73.4 |
| 622 | 12 | 82 | 65 | 77 | 74.7 | 62.1 | 82.3 |
| 403 | 10.9 | 94 | 75 | 58 | 89.8 | 72.6 | 64.9 |
| 395 | 11.7 | 18 | 75 | 38 | 18.9 | 73.4 | 36.9 |
| 388 | 12.2 | 54 | 59 | 82 | 55.8 | 63.1 | 83 |
| 172 | 10.3 | 25 | 64 | 75 | 26.9 | 62.3 | 77 |
| 137 | 13.3 | 34 | 101 | 39 | 33.4 | 101 | 37.6 |
| 123 | 11.2 | 34 | 62 | 57 | 32.3 | 65.2 | 55.6 |
| 119 | 9.59 | 54 | 44 | 14 | 53.9 | 46.1 | 15.9 |
| 107 | 9.81 | 87 | 60 | 77 | 86.5 | 61.2 | 77.1 |
| 76 | 10.5 | 80 | 99 | 37 | 79.5 | 99.7 | 36.8 |
| 54 | 9.59 | 27 | 89 | 25 | 27.9 | 88.3 | 25.5 |
| 45 | 9.89 | 78 | 65 | 56 | 78.1 | 62.8 | 55.9 |
| 43 | 10.2 | 48 | 25 | 21 | 48 | 25.8 | 20.7 |
| 42 | 10.3 | 57 | 100 | 50 | 57.6 | 98.8 | 51.3 |
| 41 | 9.4 | 48 | 54 | 88 | 48.3 | 52.9 | 87.2 |
| 29 | 9.36 | 81 | 33 | 22 | 80.6 | 33.5 | 22.2 |
| 27 | 11.3 | 52 | 97 | 54 | 52.8 | 98 | 52 |
| 26 | 8.67 | 91 | 50 | 38 | 91 | 51.2 | 37.8 |
| 21 | 9.64 | 31 | 34 | 22 | 31.5 | 34.3 | 22 |
| 21 | 9.93 | 58 | 82 | 45 | 58.4 | 82.8 | 44.4 |
| 21 | 9.44 | 81 | 58 | 52 | 80.7 | 57.6 | 53.1 |
| 18 | 8.53 | 85 | 93 | 59 | 85.4 | 93.1 | 59.4 |
| 17 | 8.76 | 79 | 54 | 82 | 77.2 | 54.3 | 82.2 |
| 17 | 9.13 | 39 | 89 | 34 | 37.9 | 89.4 | 34.5 |
| 15 | 8.87 | 53 | 81 | 63 | 52.6 | 81 | 63.2 |
| 15 | 8.15 | 74 | 87 | 34 | 73.8 | 88.5 | 34.3 |
| 13 | 8.61 | 87 | 64 | 41 | 86.5 | 61.5 | 41.7 |
| 11 | 8.5 | 34 | 71 | 87 | 35.4 | 71.9 | 85.3 |

Table 13: Coordinates of the functional connectivity map to the default mode network meta-analytical cluster 7 (thalamus), at threshold $t > 7$, in the HCP 7T dataset.

| Number of voxels | Peak t statistic | Peak X (voxel) | Peak Y (voxel) | Peak Z (voxel) | COG X (voxel) | COG Y (voxel) | COG Z (voxel) |
|---|---|---|---|---|---|---|---|
| 5659 | 19.1 | 58 | 52 | 58 | 56.2 | 46.5 | 63.7 |
| 2873 | 13.7 | 56 | 102 | 55 | 56.3 | 108 | 51.1 |
| 2174 | 85.5 | 56 | 71 | 50 | 56.2 | 69.9 | 49.7 |
| 473 | 10.2 | 65 | 15 | 42 | 62.9 | 19.5 | 42.2 |
| 289 | 11 | 49 | 18 | 41 | 45.6 | 17.9 | 43.2 |
| 266 | 10.5 | 85 | 33 | 65 | 83.1 | 34.3 | 67.6 |
| 250 | 9.73 | 28 | 38 | 67 | 25.2 | 40.7 | 64.5 |
| 219 | 13.2 | 58 | 46 | 23 | 60.3 | 47.7 | 20.4 |
| 156 | 11.3 | 49 | 48 | 20 | 48.9 | 48.6 | 16.9 |
| 104 | 11.9 | 78 | 32 | 23 | 79.6 | 33.2 | 22.1 |
| 104 | 9.2 | 72 | 97 | 73 | 69.8 | 97.2 | 72 |
| 60 | 13.5 | 56 | 66 | 33 | 56.1 | 67.7 | 33.9 |
| 53 | 9.4 | 41 | 98 | 75 | 41.9 | 96.4 | 74.2 |
| 53 | 8.69 | 89 | 39 | 66 | 88 | 39.6 | 64.8 |
| 46 | 9.17 | 30 | 35 | 22 | 33.2 | 32.9 | 22.4 |
| 38 | 09.07 | 64 | 27 | 19 | 65.3 | 26.1 | 19.7 |
| 31 | 9.98 | 33 | 101 | 38 | 33.6 | 101 | 38.2 |
| 29 | 8.99 | 49 | 25 | 21 | 48.2 | 25.9 | 20.8 |
| 16 | 09.03 | 42 | 31 | 30 | 41.6 | 31.8 | 29.4 |
| 14 | 7.95 | 41 | 103 | 70 | 39.9 | 102 | 69.4 |
| 13 | 9.43 | 56 | 86 | 43 | 56.3 | 88.1 | 43.2 |

Table 14: Coordinates of the functional connectivity map to the default mode network meta-analytical cluster 8 (Cerebellum R), at threshold t > 7, in the HCP 7T dataset.

| Number of voxels | Peak t statistic | Peak X (voxel) | Peak Y (voxel) | Peak Z (voxel) | COG X (voxel) | COG Y (voxel) | COG Z (voxel) |
|---|---|---|---|---|---|---|---|
| 7133 | 18.6 | 58 | 110 | 67 | 59.4 | 107 | 62.5 |
| 3893 | 22 | 56 | 46 | 67 | 56.7 | 44.1 | 64.6 |
| 3548 | 17.5 | 93 | 55 | 45 | 90.6 | 69.6 | 35.1 |
| 2876 | 51.4 | 37 | 30 | 25 | 38.4 | 30.6 | 23.6 |
| 2185 | 21 | 91 | 39 | 64 | 86.6 | 40.4 | 63.6 |
| 1669 | 14.1 | 29 | 87 | 25 | 21.8 | 75.7 | 32.2 |
| 1509 | 19.8 | 75 | 30 | 24 | 73 | 29.4 | 23.2 |
| 1132 | 16.7 | 91 | 94 | 52 | 84.8 | 95.9 | 42.7 |
| 918 | 17.6 | 25 | 42 | 64 | 24 | 42.6 | 62.8 |
| 724 | 19.7 | 52 | 42 | 18 | 55.1 | 44.8 | 17.7 |
| 391 | 11.4 | 24 | 100 | 37 | 29.8 | 97 | 37.1 |
| 187 | 15.5 | 57 | 70 | 68 | 56.3 | 67.4 | 68.8 |
| 77 | 10.7 | 60 | 59 | 48 | 59.7 | 60.8 | 48.7 |
| 64 | 10.2 | 51 | 81 | 55 | 51 | 85.5 | 52.9 |
| 54 | 11.3 | 61 | 86 | 53 | 61.6 | 84.9 | 53.4 |
| 37 | 9.54 | 20 | 96 | 51 | 21.3 | 95.5 | 51.1 |
| 27 | 9.82 | 72 | 66 | 35 | 70.7 | 65.7 | 35.6 |
| 27 | 8.96 | 57 | 103 | 31 | 56.3 | 103 | 31.2 |
| 15 | 8.68 | 40 | 67 | 35 | 40.9 | 66.7 | 35.3 |
| 14 | 9.28 | 72 | 116 | 45 | 71.6 | 116 | 43.9 |
| 11 | 8.23 | 51 | 61 | 51 | 50.8 | 60.7 | 50.4 |

Table 15: Coordinates of the functional connectivity map to the default mode network meta-analytical cluster 9 (cerebellum), at threshold t $> 7$, in the HCP 7T dataset.

| Number of voxels | Peak t statistic | Peak X (voxel) | Peak Y (voxel) | Peak Z (voxel) | COG X (voxel) | COG Y (voxel) | COG Z (voxel) |
|---|---|---|---|---|---|---|---|
| 10602 | 18.8 | 41 | 97 | 76 | 54.1 | 105 | 61.4 |
| 8251 | 25.3 | 56 | 38 | 66 | 55.9 | 44.9 | 63.2 |
| 2641 | 18.7 | 78 | 32 | 23 | 73.6 | 31 | 22.7 |
| 2503 | 18.9 | 88 | 35 | 67 | 84.4 | 38.2 | 65.4 |
| 2418 | 22 | 23 | 41 | 63 | 25.4 | 41.1 | 64.5 |
| 2136 | 14.9 | 17 | 75 | 38 | 19.8 | 71.8 | 35.1 |
| 2096 | 14.9 | 94 | 72 | 39 | 92.3 | 69 | 36.3 |
| 1759 | 16.2 | 31 | 35 | 22 | 38 | 31.2 | 23.2 |
| 1195 | 70.5 | 61 | 44 | 17 | 57.1 | 45.4 | 17 |
| 422 | 14.7 | 34 | 102 | 39 | 31 | 99.5 | 36.7 |
| 303 | 14.7 | 42 | 66 | 36 | 40.9 | 63.9 | 33.5 |
| 234 | 12.6 | 72 | 57 | 34 | 71.8 | 56.6 | 35 |
| 147 | 15.2 | 71 | 66 | 35 | 69.5 | 67.4 | 33.2 |
| 117 | 12 | 50 | 61 | 51 | 51.8 | 64 | 50.7 |
| 112 | 12.5 | 51 | 84 | 54 | 49.6 | 85.5 | 53.9 |
| 98 | 10.03 | 62 | 61 | 51 | 60 | 62.5 | 50 |
| 83 | 11.8 | 61 | 86 | 53 | 60.9 | 86.1 | 50.8 |
| 42 | 13.9 | 56 | 65 | 32 | 55.9 | 66.2 | 32.8 |
| 39 | 8.57 | 20 | 94 | 55 | 20.7 | 95.4 | 54.2 |
| 27 | 9.92 | 74 | 88 | 34 | 74.3 | 88.7 | 34.3 |
| 24 | 8.3 | 47 | 117 | 40 | 48.8 | 118 | 38.6 |
| 17 | 10.6 | 53 | 87 | 43 | 52.8 | 87.5 | 43.7 |
| 17 | 8.43 | 84 | 90 | 63 | 84.2 | 89.7 | 63.9 |
| 16 | 8.24 | 56 | 103 | 32 | 55.9 | 103 | 31.6 |
| 16 | 9.43 | 56 | 86 | 42 | 55.5 | 87.9 | 41.7 |
| 14 | 8.37 | 89 | 92 | 53 | 90.2 | 93.8 | 51.3 |
| 12 | 8.2 | 88 | 101 | 37 | 87.1 | 101 | 37.5 |
| 12 | 8.23 | 32 | 114 | 39 | 31.9 | 114 | 39.9 |

Table 16: Coordinates of the functional connectivity map to the default mode network meta-analytical cluster 10 (MFG, SFG), at threshold t > 7, in the HCP 7T dataset.

| Number of voxels | Peak t statistic | Peak X (voxel) | Peak Y (voxel) | Peak Z (voxel) | COG X (voxel) | COG Y (voxel) | COG Z (voxel) |
|---|---|---|---|---|---|---|---|
| 13654 | 35.2 | 54 | 110 | 66 | 60.6 | 105 | 60.6 |
| 5958 | 24.5 | 56 | 42 | 64 | 56.4 | 45.7 | 64.2 |
| 3830 | 19.2 | 95 | 57 | 45 | 90.8 | 69.2 | 35.3 |
| 2885 | 24.8 | 91 | 40 | 62 | 86.5 | 40.5 | 63.9 |
| 2739 | 17.9 | 15 | 61 | 43 | 21.2 | 72.5 | 34.3 |
| 2166 | 17.1 | 37 | 31 | 24 | 39.5 | 29.8 | 24.1 |
| 2047 | 21.8 | 24 | 42 | 64 | 23.7 | 43.3 | 63 |
| 1620 | 17.9 | 77 | 31 | 23 | 72.7 | 29.4 | 23.5 |
| 1380 | 17.3 | 38 | 91 | 36 | 28.5 | 97.2 | 40.5 |
| 816 | 19.5 | 53 | 43 | 18 | 55.6 | 45 | 17.8 |
| 154 | 12.1 | 50 | 87 | 54 | 49.8 | 84.7 | 53.9 |
| 154 | 12.3 | 61 | 86 | 53 | 62.4 | 83.4 | 54.3 |
| 81 | 11.2 | 62 | 60 | 50 | 60.4 | 60.6 | 47.8 |
| 58 | 11.6 | 71 | 65 | 36 | 71.7 | 65.7 | 34.9 |
| 53 | 12 | 39 | 65 | 36 | 39.2 | 67.1 | 34.4 |
| 45 | 10.6 | 51 | 62 | 51 | 51 | 61.1 | 49.2 |
| 40 | 9.19 | 72 | 58 | 34 | 72.3 | 56.1 | 35.2 |
| 36 | 9.52 | 58 | 66 | 52 | 58 | 67.4 | 51 |
| 29 | 10.1 | 33 | 84 | 21 | 33.5 | 85.5 | 20.4 |
| 27 | 9.05 | 57 | 103 | 31 | 56.5 | 103 | 31.2 |
| 18 | 11.7 | 56 | 67 | 32 | 56 | 66.7 | 32.2 |

Table 17: Coordinates of the functional connectivity map to the default mode network meta-analytical cluster 10 (MFG, SFG), at threshold t > 7, in the HCP 7T dataset.

| Number of voxels | COG X (voxel) | COG Y (voxel) | COG Z (voxel) |
|---|---|---|---|
| 8739 | 57.7 | 108 | 57.3 |
| 6274 | 56.6 | 44.6 | 63.5 |
| 1901 | 84.3 | 38.2 | 65.3 |
| 1399 | 24.6 | 41.9 | 63.3 |
| 742 | 94 | 71.2 | 35.5 |
| 638 | 55.7 | 45.9 | 17 |
| 324 | 18.4 | 74.4 | 35.2 |
| 317 | 29.7 | 35.6 | 21.2 |
| 280 | 33.7 | 96.8 | 35.7 |
| 174 | 81.3 | 33.9 | 21.9 |
| 173 | 70.5 | 64.7 | 35.1 |
| 152 | 64.4 | 25.6 | 20.1 |
| 148 | 47.8 | 25.3 | 21.2 |
| 105 | 40.3 | 66.2 | 35.2 |
| 105 | 55.9 | 86.4 | 42.5 |
| 74 | 74.7 | 88.2 | 34.3 |
| 71 | 78.6 | 98.2 | 35.4 |
| 55 | 71.8 | 56.9 | 34.9 |
| 49 | 61.4 | 60.2 | 47.5 |
| 38 | 57.1 | 72.5 | 67.2 |
| 37 | 29.7 | 86.7 | 75.7 |
| 33 | 50.7 | 61 | 48.8 |
| 28 | 56.3 | 65.1 | 32 |
| 19 | 49.5 | 90.5 | 51.3 |
| 14 | 58.6 | 78.9 | 44.6 |
| 14 | 44.3 | 71.6 | 26.4 |

### 3 Supplementary figures

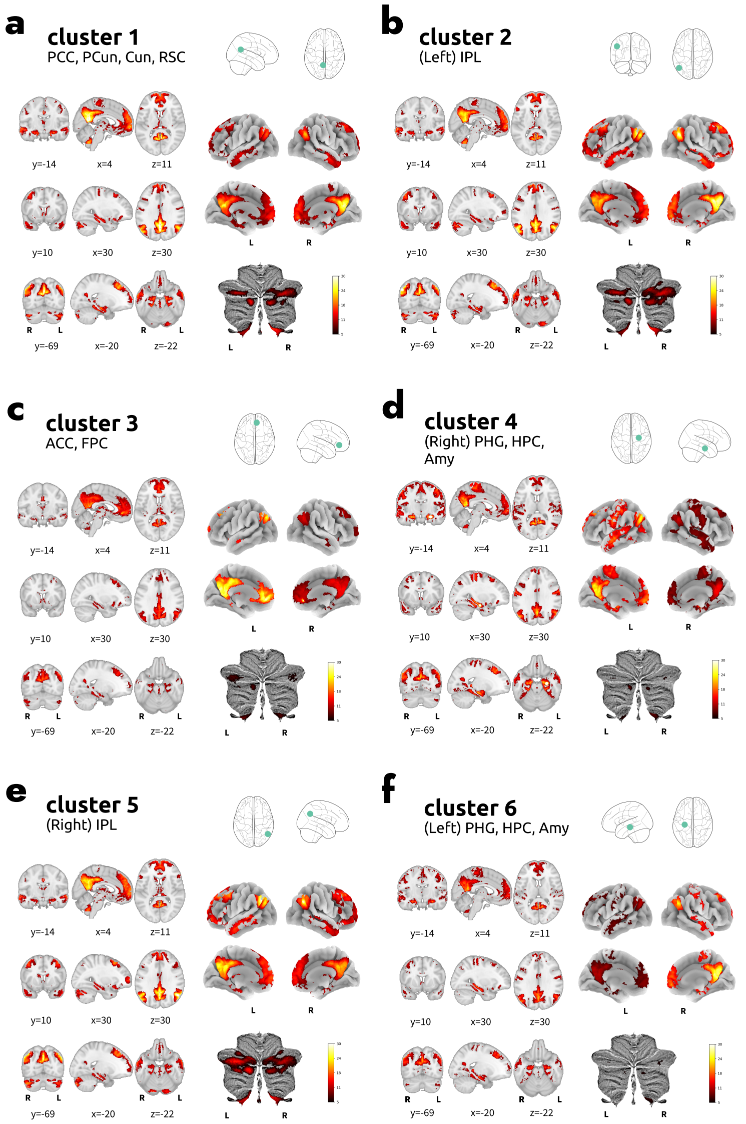


Figure 1: Functional connectivity maps from each default mode network ROI. From a to f, functional connectivity maps (t values) of clusters 1 to 6.

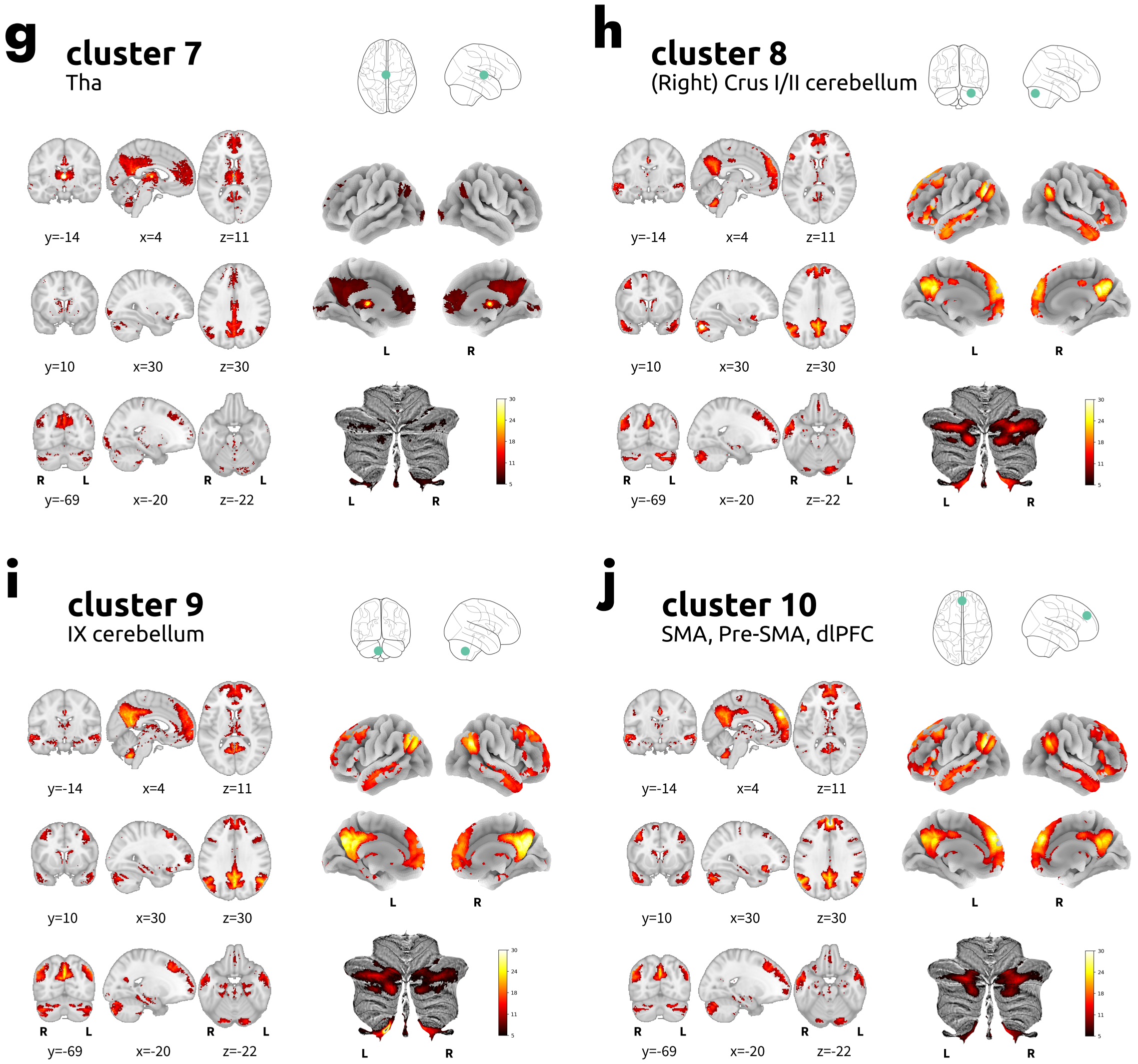


Figure 2: Continuation of functional connectivity maps from each default mode network ROI. From g to j, functional connectivity maps (t values) of clusters 7 to 10.

[26] L. Griffanti, G. Salimi-Khorshidi, C. F. Beckmann, E. J. Auerbach, G. Douaud, C. E. Sexton, E. Zsoldos, K. P. Ebmeier, N. Filippini, C. E. Mackay, S. Moeller, J. Xu, E. Yacoub, G. Baselli, K. Ugurbil, K. L. Miller, and S. M. Smith. ICA-based

artefact removal and accelerated fMRI acquisition for improved resting state network imaging. *Neuroimage*, 95:232–247, July 2014.

[27] O. Grigg and C. L. Grady. The default network and processing of personally relevant information: converging evidence from task-related modulations and functional connectivity. *Neuropsychologia*, 48(13):3815–3823, Nov. 2010.

[28] E. Gultepe and B. He. A linear/nonlinear characterization of resting state brain networks in FMRI time series. *Brain Topogr.*, 26(1):39–49, Jan. 2013.

[29] A. Hafkemeijer, C. Möller, E. G. P. Dopper, L. C. Jiskoot, T. M. Schouten, J. C. van Swieten, W. M. van der Flier, H. Vrenken, Y. A. L. Pijnenburg, F. Barkhof, P. Scheltens, J. van der Grond, and S. A. R. B. Rombouts. Resting state functional connectivity differences between behavioral variant frontotemporal dementia and alzheimer's disease. *Front. Hum. Neurosci.*, 9:474, Sept. 2015.

[30] M. Herdick, M. Dyrba, H.-C. J. Fritz, S. Altenstein, T. Ballarini, F. Brosseron, K. Buerger, A. Can Cetindag, P. Dechent, L. Dobisch, E. Duezel, B. Ertl-Wagner, K. Fliessbach, S. Dawn Freiesleben, I. Frommann, W. Glanz, J. Dylan Haynes, M. T. Heneka, D. Janowitz, I. Kilimann, C. Laske, C. D. Metzger, M. H. Munk, O. Peters, J. Priller, N. Roy, K. Scheffler, A. Schneider, A. Spottke, E. Jakob Spruth, M. Tscheuschler, R. Vukovich, J. Wiltfang, F. Jessen, S. Teipel, and M. J. Grothe. Multimodal MRI analysis of basal forebrain structure and function across the alzheimer's disease spectrum. *Neuroimage Clin*, 28:102495, Nov. 2020.

[64] H. Zhang, S. Wang, J. Xing, B. Liu, Z. Ma, M. Yang, Z. Zhang, and G. Teng. Detection of PCC functional connectivity characteristics in resting-state fMRI in mild alzheimer's disease, 2009.

[65] W. Zheng, X. Liu, H. Song, K. Li, and Z. Wang. Altered functional connectivity of Cognitive-Related cerebellar subregions in alzheimer's disease. *Front. Aging Neurosci.*, 9:143, May 2017.

[66] J. Zhou, M. D. Greicius, E. D. Gennatas, M. E. Growdon, J. Y. Jang, G. D. Rabinovici, J. H. Kramer, M. Weiner, B. L. Miller, and W. W. Seeley. Divergent network connectivity changes in behavioural variant frontotemporal dementia and alzheimer's disease. *Brain*, 133(Pt 5):1352–1367, May 2010.

[67] Z. Zhou, X. Wang, N. J. Klahr, W. Liu, D. Arias, H. Liu, K. M. von Deneen, Y. Wen, Z. Lu, D. Xu, and Y. Liu. A conditional granger causality model approach for group analysis in functional magnetic resonance imaging. *Magn. Reson. Imaging*, 29(3): 418–433, Apr. 2011.

[68] Z. Zhou, R. Zhu, W. Shao, S.-j. Zhang, L. Wang, X.-j. Ding, and D.-t. Peng. Changes in resting-state functional connectivity of cerebellum in amnestic mild cognitive impairment and alzheimer's disease: A case-control study. *Frontiers in systems neuroscience*, 15:596221, 2021.
